# MRSD: a novel quantitative approach for assessing suitability of RNA-seq in the clinical investigation of mis-splicing in Mendelian disease

**DOI:** 10.1101/2021.03.19.21253973

**Authors:** Charlie F. Rowlands, Algy Taylor, Gillian Rice, Nicola Whiffin, Hildegard Nikki Hall, William G. Newman, Graeme C.M. Black, kConFab Investigators, Raymond T. O’Keefe, Simon Hubbard, Andrew G.L. Douglas, Diana Baralle, Tracy A. Briggs, Jamie M. Ellingford

## Abstract

**Background:** RNA-sequencing of patient biosamples is a promising approach to delineate the impact of genomic variants on splicing, but variable gene expression between tissues complicates selection of appropriate tissues. Relative expression level is often used as a metric to predict RNA-sequencing utility. Here, we describe a gene- and tissue-specific metric to inform the feasibility of RNA-sequencing, overcoming some issues with using expression values alone.

**Results:** We derive a novel metric, *Minimum Required Sequencing Depth* (MRSD), for all genes across three human biosamples (whole blood, lymphoblastoid cell lines (LCLs) and skeletal muscle). MRSD estimates the depth of sequencing required from RNA-sequencing to achieve user-specified sequencing coverage of a gene, transcript or group of genes of interest. MRSD predicts levels of splice junction coverage with high precision (90.1-98.2%) and overcomes transcript region-specific sequencing biases. Applying MRSD scoring to established disease gene panels shows that LCLs are the optimum source of RNA, of the three investigated biosamples, for 69.3% of gene panels. Our approach demonstrates that up to 59.4% of variants of uncertain significance in ClinVar predicted to impact splicing could be functionally assayed by RNA-sequencing in at least one of the investigated biosamples.

**Conclusions:** We demonstrate the power of MRSD as a metric to inform choice of appropriate biosamples for the functional assessment of splicing aberrations. We apply MRSD in the context of Mendelian genetic disorders and illustrate its benefits over expression-based approaches. We anticipate that the integration of MRSD into clinical pipelines will improve variant interpretation and, ultimately, diagnostic yield.

## Introduction

Pinpointing disease-causing genomic variation informs diagnosis, treatment and management for a wide range of rare disorders. An underappreciated group of pathogenic variants is those that lie outside of canonical splice sites but act through disruption of pre-mRNA splicing, the process whereby introns are removed from nascent pre-mRNA to produce mature and functional transcripts (Supplementary Figure 1a). The ways through which genomic variants can disrupt pre-mRNA splicing are diverse (Supplementary Figures 1b-g), including both protein-coding and intronic variants that are well described as causes of rare disorders (1-3). However, the omission of intronic regions in targeted sequencing approaches (4, 5), discordance between *in silico* variant prioritization tools (6) and the lack of availability of the appropriate tissue from which to survey RNA for splicing disruption (7, 8) limit effective identification of pathogenic splice-impacting variants.

RNA sequencing (RNA-seq) offers a potential route to overcome issues of variant interpretation (3, 9-12). The complex impacts of variants on splicing can be fully characterized through RNA-seq. Moreover, aberrant splicing events can be identified from RNA-seq datasets without prior knowledge of genomic variants driving their impact. Whilst targeted analyses, such as RT-PCR, also enable detection of splicing aberrations (3), such approaches are designed to test the presence of specific disruptions and may not identify the complete spectrum of splicing disruption caused by a single genomic variant.

There is growing evidence that RNA-seq can substantially improve diagnostic yield across a variety of disease subtypes (3, 10, 13-15) through identification of variants impacting splicing, or leading to impairment of transcript expression or stability (16). However, there remain several hurdles to the effective and routine integration of RNA-seq into diagnostic pipelines. For example, surveying a whole transcriptome identifies a large number of aberrant splicing events – in the order of hundreds of thousands – and there is little consensus regarding the best approach to filter for true positive and pathogenic events. Furthermore, diagnostic analysis using RNA-seq is only effective when sufficient levels of sequence coverage of a relevant gene transcript are present in the sampled tissue.

In this study, we develop an informatics approach to quantify the likelihood that a gene/transcript, or a defined set of genes or transcripts, can be appropriately surveyed using RNA-seq. We name our framework the *Minimum Required Sequencing Depth* (MRSD), which can be utilized in a flexible and customized manner to assess the suitability of RNA-seq derived from different tissues to identify pathogenic splicing aberrations in specific genes of interest. MRSD scores (available at: https://mcgm-mrsd.github.io/) can be utilized to select the most appropriate biosample to detect splicing aberrations for a candidate set of genes/transcripts or to guide the amount of sequencing reads from a specific biosample required to generate appropriate transcriptomic datasets for a gene of interest. We apply these techniques to the study of monogenic disease genes, and assess three clinically accessible biosamples for their appropriateness to survey all known monogenic disease genes.

## Results

### Minimum Required Sequencing Depth scores differ across biosamples

We first derived MRSD scores, corresponding to the required sequencing depth (in M uniquely mapping sequencing reads) for a specified level of coverage of a transcript, for 3112 known multi-exon disease genes in three distinct tissues (blood, LCLs and skeletal muscle). Three parameters can be altered for the MRSD model; we observed that MRSDs differed dependent on the values chosen for these parameters, comprising the number of reads desired to cover each splice junction, the proportion of splicing junctions for each gene that meet this coverage threshold (75% or 95%), and the proportion of samples for which the prediction is predicted to be sufficient (the “confidence level” of either 95% or 99%; Figure 1). For example, across all three tissues at a specified read coverage level of eight reads per splicing junction, we observed that increases in the desired proportion of covered splice junctions from 75-95% was associated with an increase in median MRSD of between 5.4% (in blood) to 61.2% (in LCLs; Figure 1a, top). In general, increasing desired confidence level for appropriate splice junction coverage from 95% to 99% resulted in an increase in median MRSD of between 25.8-85.8%. Conversely, for skeletal muscle samples, when stipulating 95% splice junction coverage, we observed a decrease of 3.1% in MRSD scores when desired confidence level was increased from 95% (*n* = 1241, median = 41.83) to 99% (*n* = 921, median = 40.54); this was accounted for by an increase in the number of genes that were considered “unfeasible” for surveillance, i.e. those for which zero reads cover the given proportion of junctions (*n* unfeasible at 95% confidence = 1873, *n* unfeasible at 99% confidence = 2193). This definition of feasibility is limited by the sequencing depth of the control models on which the predictions are based. For example, no coverage of splice junctions in a particular transcript may have been observed simply due to low sequencing depth; with ultra-deep sequencing of the same sample, we may have observed coverage of splice junctions and so have been able to generate a feasible MRSD prediction.

**Figure 1.**
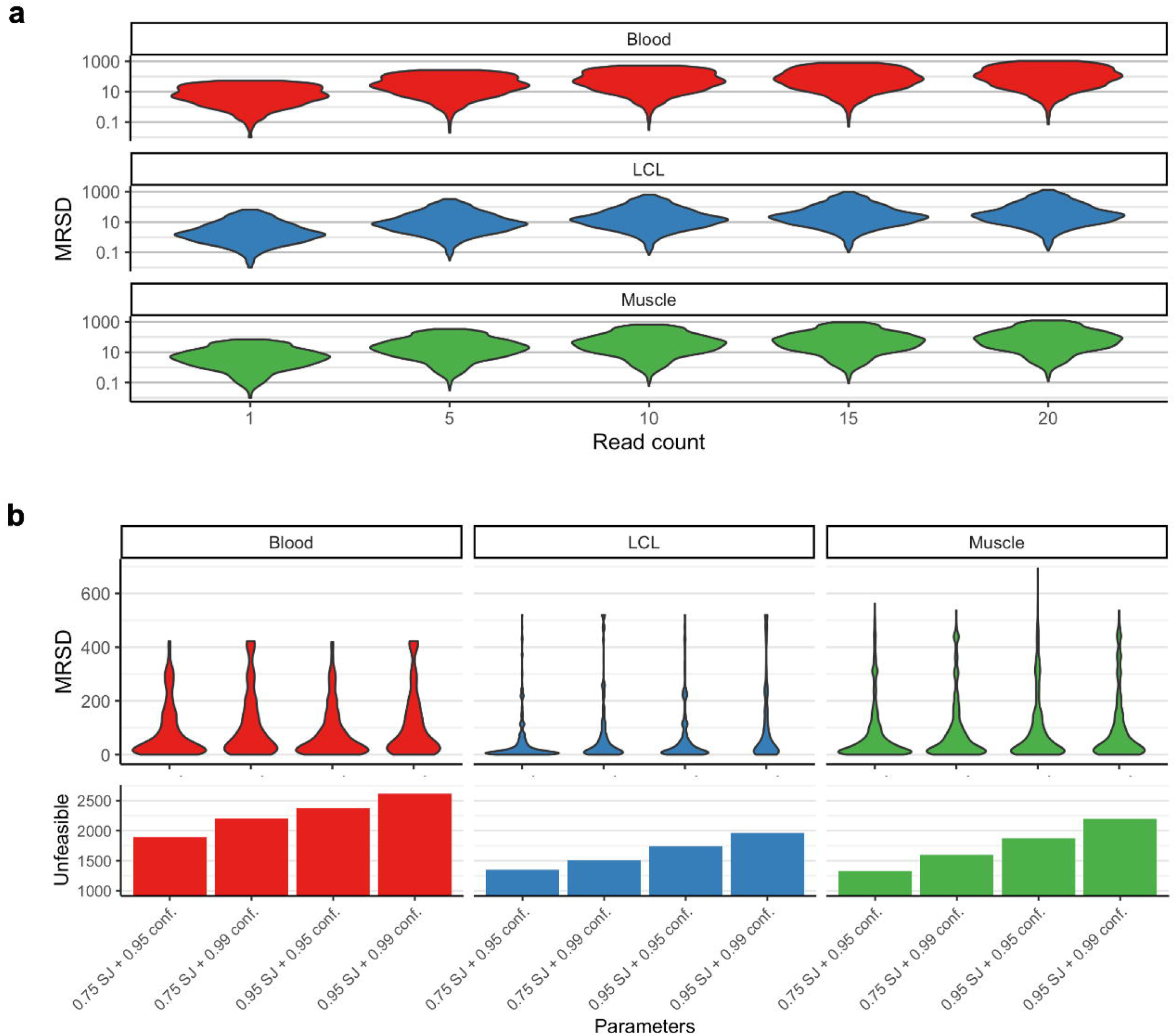
Minimum required sequencing depth (MRSD) predictions vary with changes in model parameters and across tissues. (**a**) When all other parameters are constant (default parameters used here), increasing the desired level of read coverage of a gene results in a proportional increase in MRSD. The distribution of MRSD scores for 3112 PanelApp genes in lymphoblastoid cell lines (LCLs) appears to be the lowest of the 3 tissues (median = 14.89 M at 10 reads), while whole blood exhibits the highest overall MRSD scores (median = 45.91 M at 10 reads), suggesting coverage of disease genes is generally poorer in blood. (**b**, top) In most cases, for a given level of splice junction (SJ) coverage, increasing the desired confidence level (the proportion of RNA-seq runs for which the MRSD prediction is expected to be sufficient) results in an increase in median MRSD score. (**b**, bottom) The number of genes for which no amount of sequencing is predicted to yield the specified level of coverage increases gradually as parameter stringency increases. At the highest level of stringency, the specified coverage was predicted unfeasible for between 63.1% (1964/3112, in LCLs) and 84.1% (2616/3112, in blood) of PanelApp genes.

Overall, these analyses suggested that, of the three investigated biosamples, LCLs would enable investigation of the most comprehensive set of genes for aberrant splicing. This conclusion was supported by LCLs displaying, across all four parameter combinations, the lowest median MRSDs (range = 12.86-33.77, Figure 1b, top), and the fewest “unfeasible” genes (43-63%). On the other hand, whole blood exhibited the highest number of unfeasible genes across the different parameter combinations (61-84%).

### Accuracy of Minimum Required Sequencing Depth calculations

We next obtained RNA-seq datasets for 68 samples from the three investigated tissues (blood, *n* = 12; LCLs, *n* = 4; muscle, *n* = 52), with a wide range of sequencing depths (Supplementary Figure 2). We assessed the performance of the MRSD model against these datasets, defining the positive predictive value (PPV) of MRSD as the likelihood that appropriate sequencing coverage was obtained given that the level of sequencing depth exceeded the MRSD prediction. Conversely, the negative predictive value (NPV) was defined as the likelihood that appropriate sequencing coverage was not obtained, given that the sample did not meet the specified criteria of the MRSD prediction. Across all investigated MRSD parameters, we observed 96% PPV and 79% NPV, on average, for the 68 samples (Figure 2a). We observed a general trend that the PPV and NPV of MRSD decreased and increased, respectively, as higher levels of required coverage were imposed (Figure 2b-c). Across all parameter combinations, PPV values ranged from 90.1-98.2%, while NPV ranged from 56.4-94.7%, suggesting MRSD is a fairly conservative model that primarily returns positive results with high certainty.

**Figure 2.**
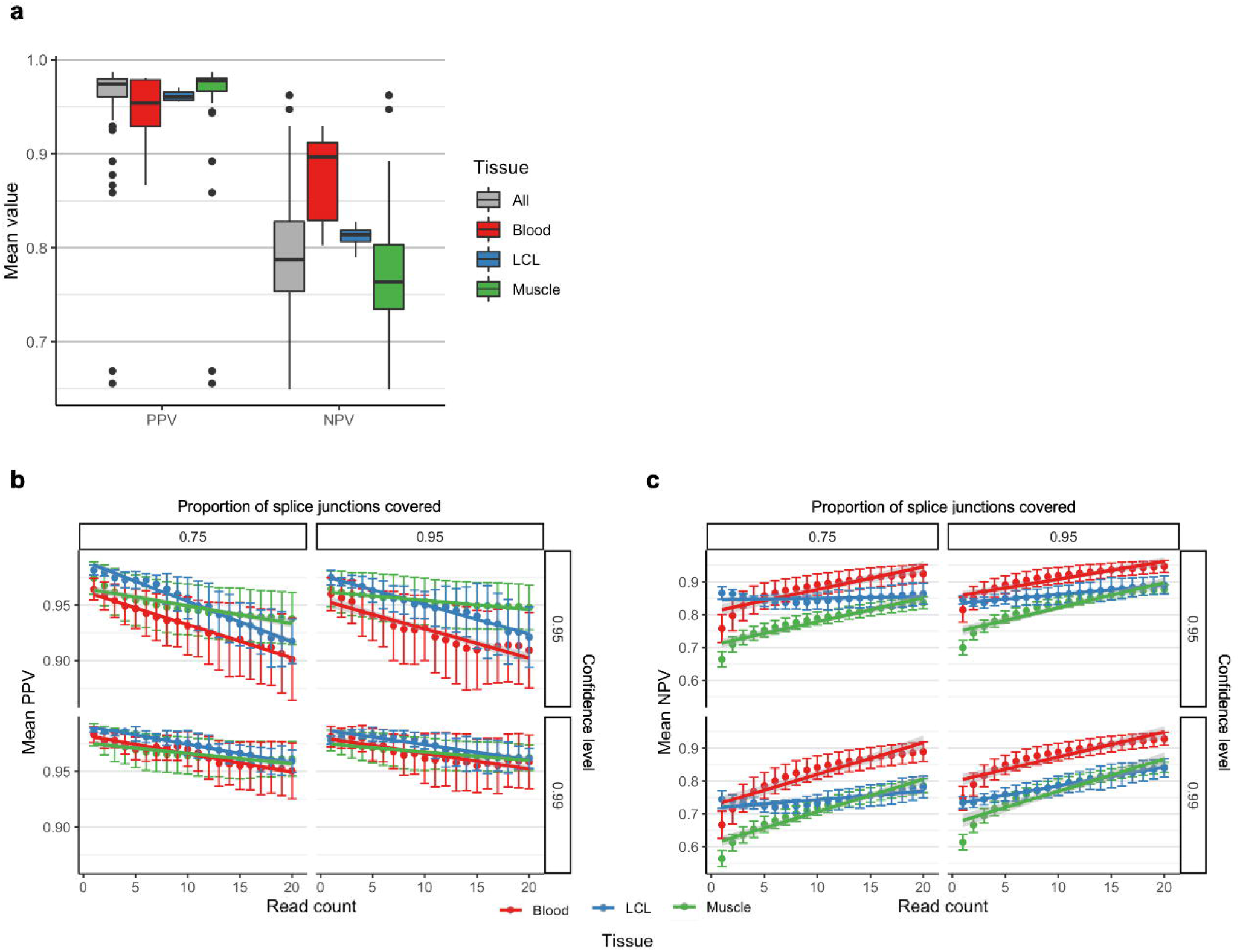
Performance metrics of the MRSD model. The ability of MRSD to accurately predict levels of PanelApp disease gene coverage based on sequencing depth was tested on unseen RNA-seq datasets from blood (*n* = 12), LCLs (*n* = 4) and muscle (*n* = 52). (**a**) The mean positive predictive values (PPVs) and negative predictive values (NPVs) averaged across all parameter combinations for each RNA-seq dataset show that the median PPV is slightly lower, and the median NPV slightly higher, for whole blood than for LCLs and skeletal muscle. Breakdown of (**b**) PPVs and (**c**) NPVs for the MRSD model by parameters shows that specifying an increasing required read coverage results in a gradual decrease in PPV and increase in NPV across all tissues and parameter combinations. Dependent on parameter stringency, and limiting analysis to a maximum specification of 20-read coverage, PPV predictions range from 90.1-98.2%, while NPV ranges from 56.4-94.7%. Overall, the model is fairly conservative and returns positive predictions only when they are deemed likely to be true.

Interestingly, although MRSD scores were derived from 75 bp paired-end RNA-seq data, evaluating the ability of the model to predict transcript coverage in 150 bp paired-end data (LCLs, *n*=20) shows higher PPV than with 75 bp data for half of the four parameter combinations tested, while NPV was only slightly lower for all combinations (Supplementary Figure 3). This suggests that, while care must be taken applying this approach to datasets derived using alternative experimental approaches, the MRSD model described here may provide a suitable approximation in the case of alternative sequencing read lengths.

### Comparison of MRSD and TPM as a guide for appropriate surveillance

We compared MRSD to the use of relative expression level (in transcripts per million, TPM) as a possible indicator of RNA-seq suitability for the detection of aberrant splicing events. We compared the expression levels, in TPM, of PanelApp genes against tissue-specific MRSD predictions, finding a negative correlation between the level of gene expression and its predicted MRSD across all three tissues (*r*^*2*^ = 0.539-0.669; Figure 3a-c). This comparison confirms that more highly-expressed genes are associated with lower MRSD scores. However, we noted significant overlap between genes grouped into low-MRSD (< 100 M reads) and high-MRSD (≥ 100 M reads) brackets. For example, among genes considered low-MRSD, TPM values ranged from 1.25-1390, while genes with high-MRSD values had TPM values between 0-4880 (Figure 3d). We quantified the overlap between these distributions, demonstrating that 98.6% of high-MRSD genes had higher TPM values than at least one low-MRSD gene. We calculated the tissue-specific median and the lowest TPM values within the low-MRSD bracket for the top 95% and 70% percentiles, and observed higher TPM values in 52.2%, 13.3% and 5.3% of high-MRSD genes, respectively (Figure 3d). The substantial overlap in the TPM values for low and high MRSD genes suggests that relative expression does not provide a wholly accurate representation of transcript coverage in RNA-seq data. Such inconsistencies may arise from bias in the regions of genes that are sequenced, for example, genes with high degrees of 3’ bias in RNA-seq datasets (Supplementary Figure 4).

**Figure 3.**
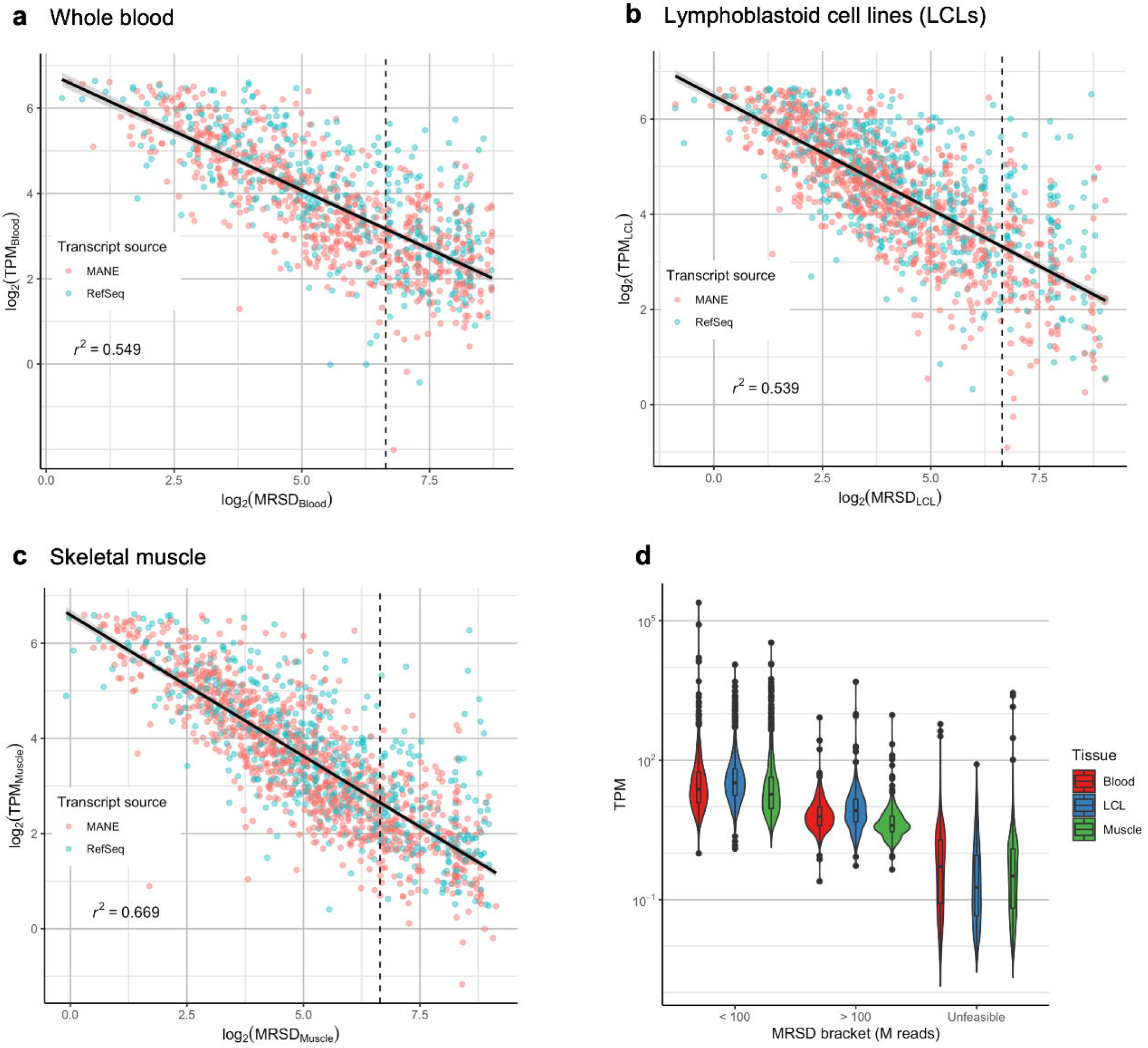
Comparison of MRSD and transcripts per million (TPM) predictions. MRSD and TPM predictions for 3112 genes present in the Genomics PanelApp repository are inversely correlated in (**a**) whole blood (*r*^2^ = 0.549), (**b**) LCLs (*r*^2^ = 0.539) and (**c**) skeletal muscle (*r*^2^ = 0.669), as might be expected; however, the correlation is broad and there is high variation in the TPMs both of genes considered low- and high-MRSD (MRSD ≤ or > 100 M reads, respectively, dotted line). (**d**) Bracketing PanelApp genes by MRSD range shows that there is substantial overlap in the TPMs of genes across different MRSD predictions, to the extent that sufficient coverage of genes with TPMs up to 2796.5 is predicted unfeasible in some cases. This suggests relative expression level alone is not an adequate proxy for transcript coverage. The y-axis is limited to 100 TPM in (**a**-**c**) for ease of visualization. Log transformation in (**d**) excludes 491 entries with TPMs of 0. Default MRSD parameters (8-read coverage of 75% of splice junctions, confidence level of 95%) used throughout.

### Traits of pathogenic splicing variation vary widely between genes and events

We identified pathogenic aberrations to splicing in 20 of the 88 samples utilizing a previously described analysis pipeline (13) with a wide variety of mis-splicing effects (Supplementary Figure 5), and calculated respective median TPM and MRSD values (Supplementary Table 1). The method for aberrant splicing detection pooled evidence for splicing junctions in reference sets to generate tissue-specific models of “healthy” splicing. We incorporated RNA-seq datasets from relevant samples into the healthy splicing models (Supplementary Table 1) and collected metrics indicative of aberrant splicing events (Box 1). We observed high variability in all metrics associated with pathogenic aberrant splicing events (Table 1). All patients harbored at least one pathogenic splicing event supported by two reads and with normalized read counts (NRCs) ≥ 0.19, and 80% of these events had a relative fold change in NRC > 19x relative to controls (Table 1). While a blanket set of parameters for all aberrant splicing events may be unsuitable, our data suggests that 90% of pathogenic events could be retained if filtering for events that were singletons (evident only in a single sample), or were non-singletons with an NRC > 0.25.

**Table 1.** Range of metrics observed for pathogenic splicing events

| Metric | Tissue |  |  |
| --- | --- | --- | --- |
|  | Whole blood (n=3) | LCLs (n=7) | Skeletal muscle (n=10) |
| Read count | 2-40 | 4-38 | 2-462 |
| NRC | 0.48-1.25 | 0.19-1.52 | 0.34-3.19 |
| NRC fold change | Singletons | 3.7-8.2 + singletons | 19.6-442 + singletons |
| Number of samples | 1 | 1-48 | 1-110 |
| Rank | 2-5 | 10-232 | 1-342 |

#### Box 1. Metrics collated during splice event analysis

- Read count – Number of split reads supporting the existence of a given splice junction
- Normalized read count (NRC) – Ratio of reads supporting a given junction compared to the adjoining canonical junction with the highest read count
- NRC fold change – fold difference in NRC for a given event between an individual and the control individual with the next-highest NRC for that event
- Number of samples – the number of individuals, across both case and controls, in which an event is present
- Rank – position of a given event in a list of significant events, when ordered by decreasing read count (for singleton events) or fold change (for non-singleton events)

### Factors influencing the likelihood of pathogenic splicing variation identification & MRSD predictions

To further define the most informative parameters for use in the MRSD model, we investigated the impact of a variety of metrics on the capability to identify pathogenic splicing events, including number of samples within the healthy reference set, the extent of read support for splicing junctions, and the relative expression of genes of interest. Overall, our analyses suggested that two supporting reads for an aberrant splicing event that is novel or has an NRC > 0.25 would reliably highlight pathogenic aberrations amongst transcriptome-wide splicing variation. These parameters are conservative and could be relaxed for the targeted investigation of variants of interest.

We first identified how the number of control samples used as a reference set for “healthy splicing” impacted our ability to identify aberrant splicing events. For all samples within our healthy splicing set, we iteratively selected groups of control samples at sizes of 30, 60 or 90. We observed that moving from 30 to 60 controls is associated with a mean reduction in event count of 19.3% (28.1% of non-singleton events, 17.1% of singleton events) across the three tissues, while increasing the control size to 90 results in a further reduction of 10.2% of events (16.5% of non-singleton events, 9.5% of singleton events; Figure 4); this effect was consistent across tissue types.

**Figure 4.**
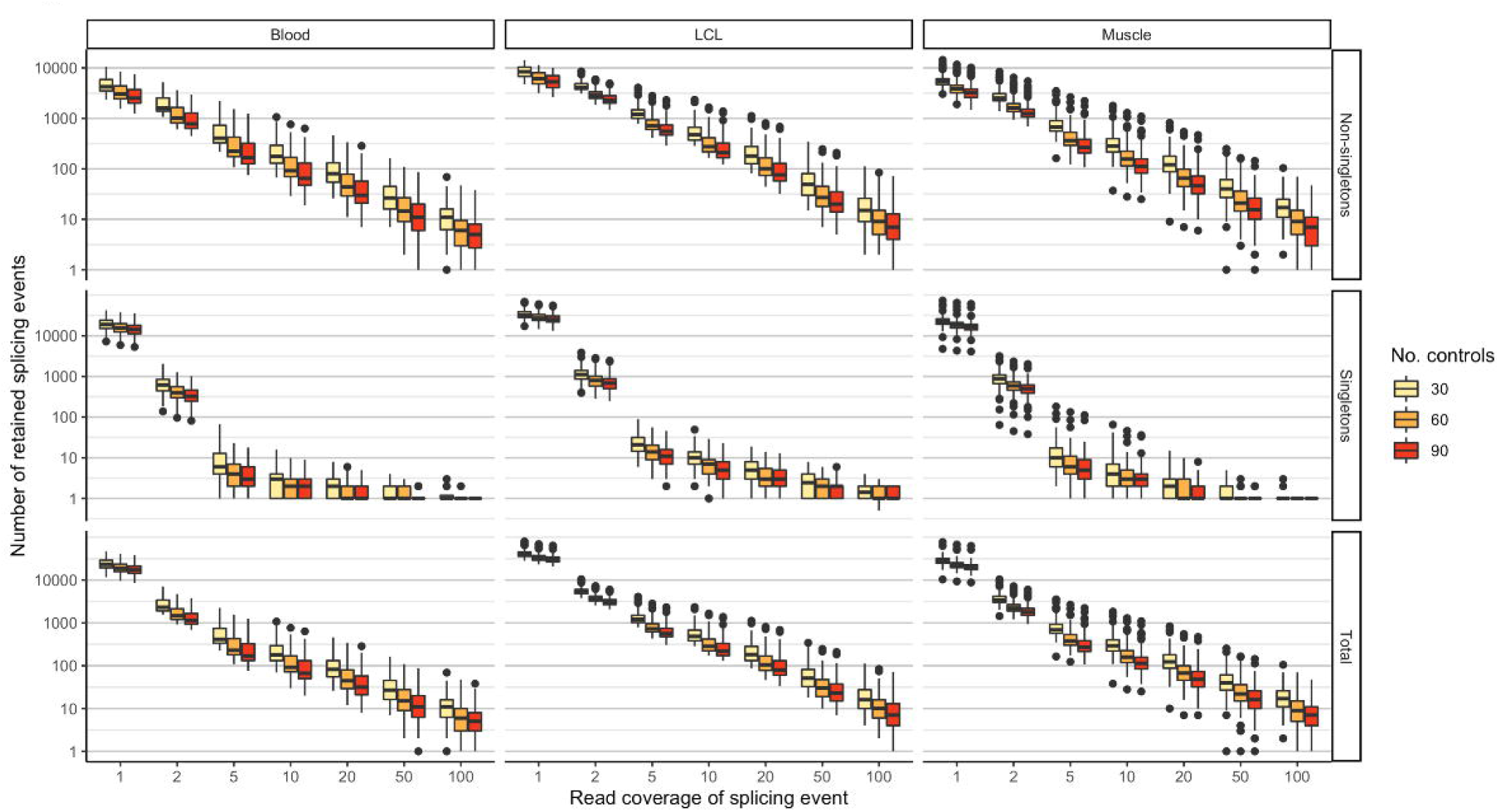
Expanding control datasets and enforcing read count thresholds improves filtering power when analyzing mis-splicing events. Counting the significant events identified in each individual in a control splicing dataset when analysed against 2000 bootstraps each of 30, 60 and 90 other individuals from within the control dataset for the same tissue reveals a small decrease in the number of total events identified as control dataset size increases, predominantly from non-singleton events. Enforcing a read coverage threshold has a more significant effect on event counts, particularly for singleton events, where filtering out events supported by a single read removes up to 95% of singletons. LCLs appear to exhibit the greatest number of splicing events regardless of filter, although this may be due to differences in sequencing depth between tissues.

We next investigated how read count filters impacted the number of events observed for a given individual (Figure 4). Filtering out all splicing events supported by just a single read against a background of 90 control samples removes, on average, 91.2% of events (60.4% of non-singleton events, 97.3% of singleton events). Increasing read support thresholds to 10 unique sequencing reads results in a total of 99.4% of events being excluded on average (96.2% of non-singleton events, 99.99% of singleton events), while retaining only those events supported by 100 reads or more removes an average of 99.97% of events (99.8% of non-singleton events, 100.0% of singleton events). To understand how the level of read support impacted the ability to identify specific events, we collated 31 aberrant splicing events across 22 muscle-derived RNA-seq samples, and downsampled reads in the genes containing these events. We observed that we could identify the same aberrant splicing events at reduced relative expression levels, and, while read support decreased (Figure 5a), the ranked position of the event within the rank-ordered output remained approximately the same in most cases (Figure 5b). However, the weakened read support increased the risk of eliminating the variant from consideration when read count filters were applied (Figure 5c). This analysis further emphasized that TPM values alone may not be a reliable measure of ability to survey all splicing junctions within a gene; we observed that splice junctions in different samples covered by the same number of sequencing reads belonged to genes with widely ranging TPM values (Supplementary Figure 6). For example, splice junctions covered by eight reads were associated with TPMs ranging between 0.17 and 52.

**Figure 5.**
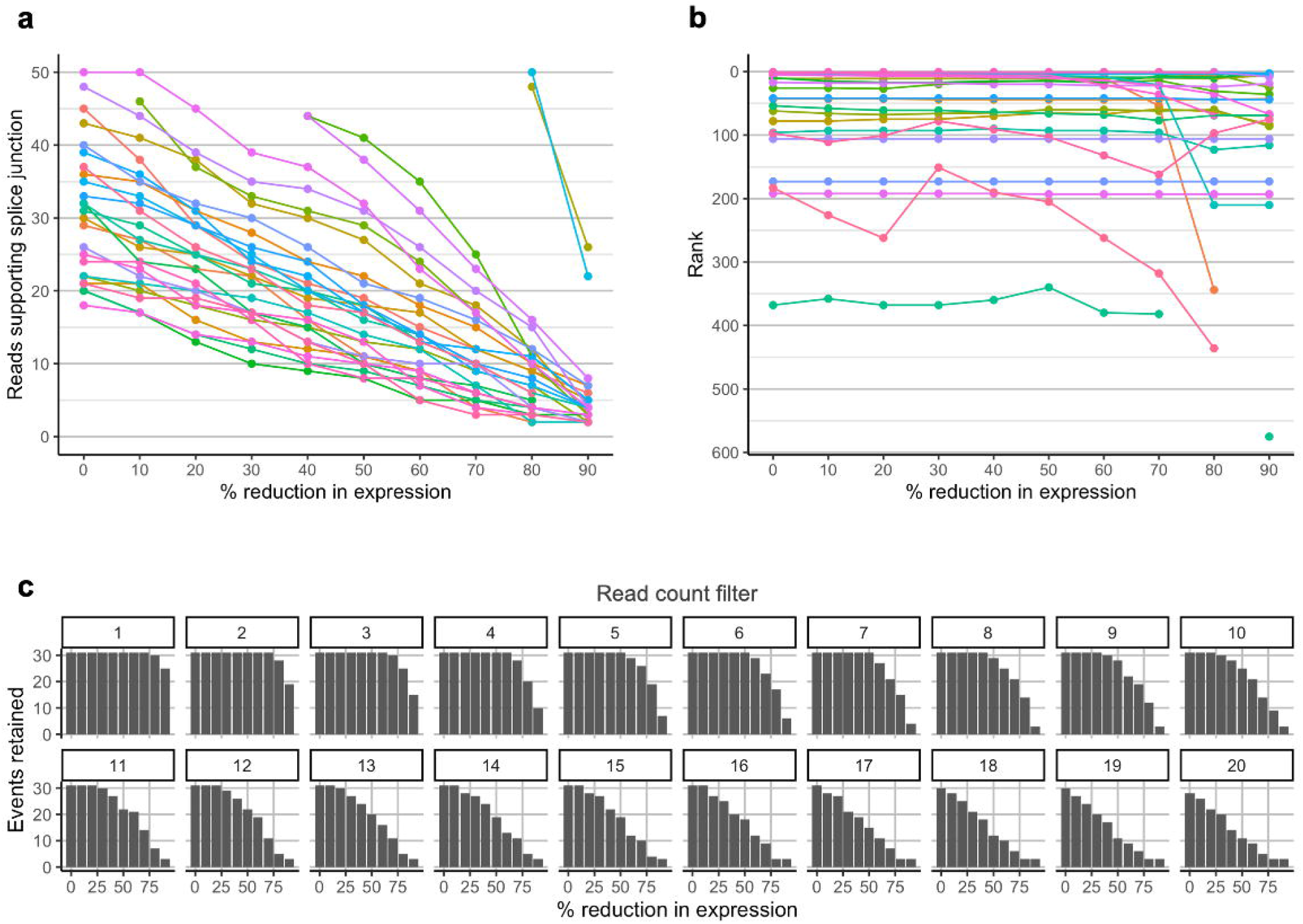
Variability in expression level influences the capacity to identify mis-splicing events. Genes harboring a selection of 31 splicing events that were identified during analysis of 52 muscle-based RNA-seq datasets (and which would be identified as events of interest using a filter of normalized read count (NRC) > 0.19) were artificially downsampled to simulate variation in expression. (**a**) Reduction in expression leads to an intuitive and proportional reduction in the number of reads supporting each mis-splicing event. (**b**) The rank position – where the event appears in a list of all splicing events in its respective sample, ordered by decreasing NRC fold change relative to controls, and – is generally consistent as expression of the gene decreases; however, for a subset of events, reduction in expression is sufficient to cause stochastic changes in the NRC value, and so cause movement of the event down the prioritized list. (**c**) Variation in expression impacts our ability to identify events of interest when filters of read count supporting the events are enforced. When the 31 events experience a 50% reduction in expression, for instance, the application of a minimum 15-read filter leads to the exclusion of 41.9% (13/31) of events. For ease of visualization, the y-axis in (**a**) is limited to 50 reads, resulting in the truncation of some data series on the graph.

### Implications for investigation of variants in known disease-causing genes

We applied our MRSD model to all established disease genes included in the Genomics England PanelApp repository, encompassing 275 distinct gene panels and 3199 unique genes. 87 single-exon genes were excluded from analysis, leaving 3112 unique disease genes. Based on our investigations of MRSD, we applied the following parameters: read coverage = 8; proportion of junctions = 75%; confidence level = 95%. Using this approach (with expected PPV = 0.936-0.974, NPV = 0.776-0.880 across the three tissues) we observed that 58.0% (1806/3112) of PanelApp genes were predicted to be low-MRSD (< 100 M reads required) in at least one of whole blood, LCLs or skeletal muscle (Figure 6a). At the individual tissue level, 27.0% (841/3112) of PanelApp genes in whole blood, 49.0% (1524/3112) in LCLs and 44.0% (1369/3112) in skeletal muscle were predicted to be low-MRSD (Figure 6a). Of note, LCLs were observed to have the highest proportion of low-MRSD panel genes in 190/275 disease-gene panels (69.3%, Figure 6c). Whole blood exhibited the highest proportion of genes with low MRSDs in just 24/275 disease-gene panels (8.8%).

**Figure 6.**
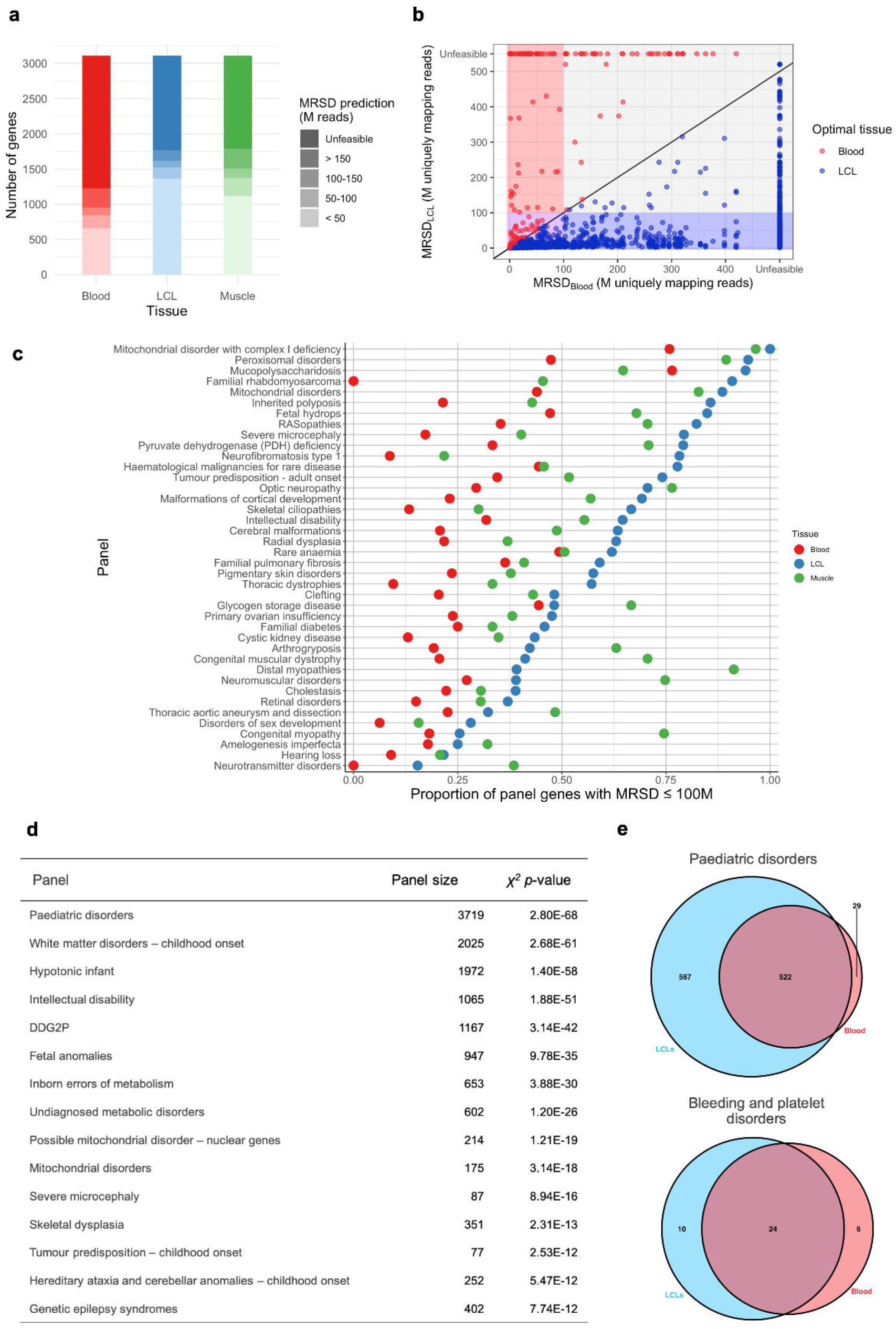
Application of MRSD scores to disease genes listed in the Genomics England PanelApp repository. (**a**) Comparison of PanelApp panel gene MRSD predictions between tissues shows blood to exhibit markedly poorer coverage of disease genes than do LCLs or skeletal muscle. (**b**) When comparing MRSD predictions for genes in blood and LCLs, 1522 genes are considered “high-MRSD” (i.e. have an MRSD > 100 M reads) in both tissues (grey). Genes which are exclusively low-MRSD (i.e. MRSD ≤ 100 M) in blood are far fewer in number (with 66 genes, red box), while the remainder are almost evenly split between those that are low-MRSD in both (775 genes, purple box) and low-MRSD in LCLs only (749 genes, blue box). (**c**) Comparison of PanelApp panel gene MRSDs between tissues shows many panel genes have substantially greater coverage in LCLs than blood and, to a lesser extent, skeletal muscle over a variety of disease subtypes. Panels where skeletal muscle shows the best coverage of panel genes intuitively correspond to phenotypes such as neuromuscular disorders and distal myopathies. 40 exemplar panels shown here, to see results for all 275 panels, see Supp. Figs. 8 & 9. (**d**) Top 10 panels with most significant difference between low- and high-MRSD gene counts between blood and LCLs (chi-squared test). (**e**) Venn diagrams showing number of low-MRSD genes predicted in blood and LCLs for (top) the paediatric disorder panel, the most significantly divergent between the two tissues, and (bottom) the bleeding and platelet disorders panel, which did not reach statistical significance in the aforementioned chi-squared analysis.

MRSD predictions revealed many use cases for specific tissues: in the familial rhabdomyosarcoma panel, for example, none of the 11 genes were predicted to be low-MRSD in blood, while 10/11 were predicted low-MRSD in LCLs (Figure 6c), of which nine were actually assigned an MRSD < 50 M reads. Results across all 275 panels are shown in Supplementary Figures 8 & 9.

Overall, this analysis suggests both that whole blood may often represent the poorest choice of RNA source tissue in terms of disease gene coverage; in contrast, LCLs appear to show robustly high expression of many disease genes across diverse disease subtypes, and so may constitute a more reliable source of RNA for clinical transcriptomic investigations.

### Quantifying the resolving power of RNA-seq for variants of uncertain significance

To analyze the possible impact of diagnostic RNA-seq integration on variant interpretation, we curated variants of uncertain significance (VUSs) from the ClinVar variant database (17) that were predicted by SpliceAI (18) to impact splicing (score ≥ 0.5; see Materials and Methods). Of a total of 352,011 ClinVar variants, 185,119 (52.6%) were identified as VUSs, and 7,507 (2.1%) were retained after filtering based on SpliceAI score. Cross-referencing the MRSDs of the genes harboring SpliceAI prioritized variants across tissues revealed that, depending on model stringency, between 22.1% and 59.4% of these variants may lie in genes that are low-MRSD in at least one of the three tissues (Figure 7a). Further, among the 30 genes in which the greatest number of predicted splice-impacting VUSs were identified, 21 were predicted to be low-MRSD in at least one tissue (Figure 7b). Similar patterns were observed when using a more relaxed SpliceAI score filter of 0.25 (Supplementary Figure 10). The guided integration of RNA-seq into diagnostic services alongside predictive bioinformatics tools is therefore likely to provide a significant improvement to interpretation of VUSs in a variety of disease contexts.

**Figure 7.**
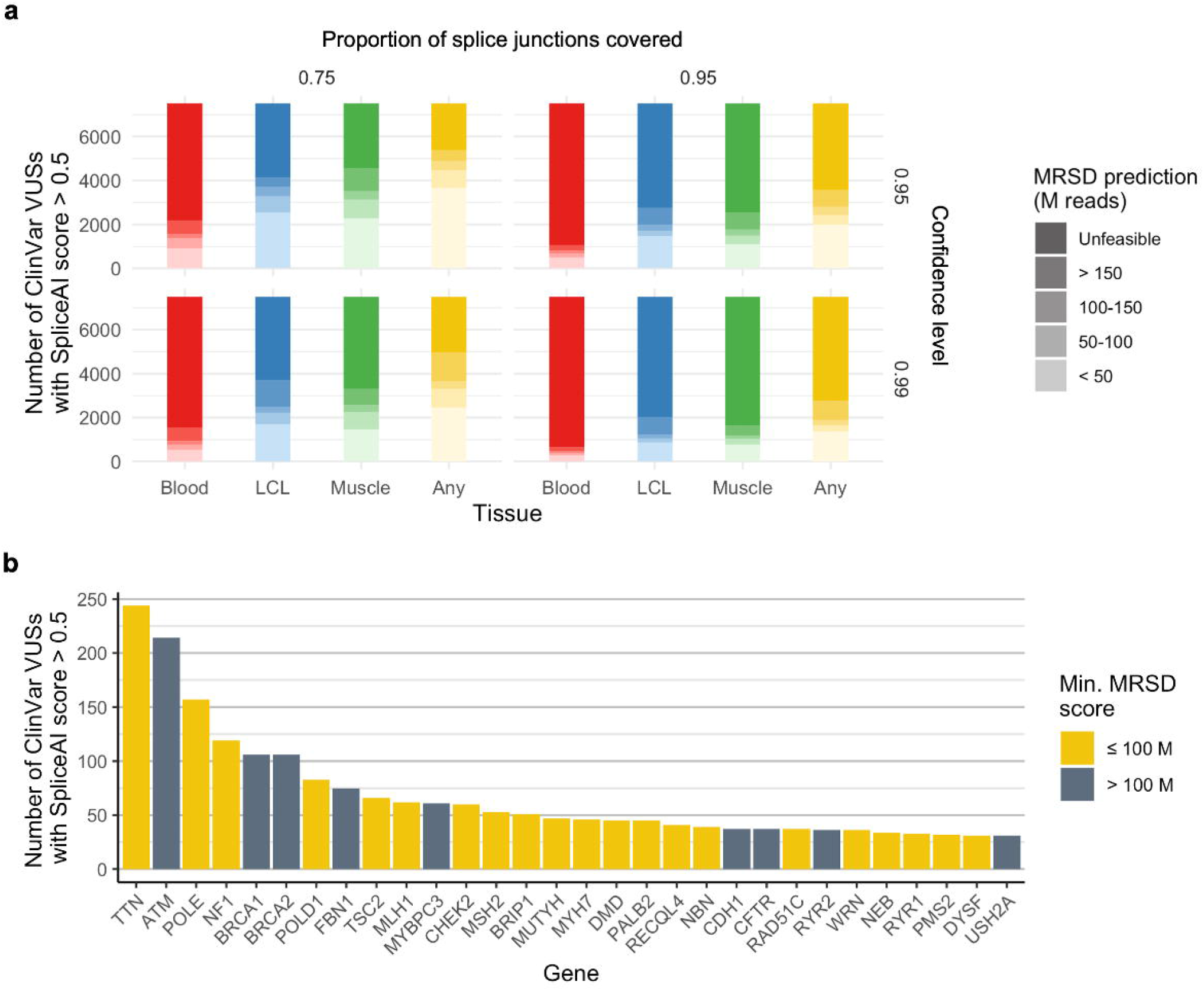
The scope for resolution of variants of uncertain significance (VUSs) using RNA-seq-based analysis. MRSD scores were derived for the genes harbouring VUSs present in ClinVar if the variants were predicted by the predictive tool SpliceAI to impact splicing (score ≥ 0.5; Jaganathan et al., 2019) (**a**) Depending on the stringency of the MRSD model parameters, between 22.1% (1663/7507) and 59.4% (4462/7507) of variants predicted to impact splicing are expected to be adequately covered by 100 M uniquely mapping reads or fewer in at least one of the 3 tissues (whole blood, LCLs and skeletal muscle). Variants were most likely to be found to be in low-MRSD genes (MRSD ≤ 100 M) in LCLs, irrespective of model parameters. (**b**) Among the 30 genes with the greatest number of predicted splice-impacting VUSs, 21 were predicted to be adequately covered (using default parameters) with 100 M uniquely mapping reads or fewer in at least one of the 3 tissues. An 8-read junction support parameter was used throughout.

## Discussion

The recent development of machine learning approaches has underpinned improvements to the prioritization of variants that impact splicing and cause rare disease (19). Despite these advances, corroboration of the effect of such variants remains a major obstacle to improving diagnostic yield for Mendelian disorders. This obstacle is amplified by the unexpected functional impact of some variants on splicing, which may change the way the variant is classified in accordance with current guidelines (6). The MRSD-based approach described here allows the informed selection of biosample(s) for bulk RNA-seq, based on the required number of sequencing reads that need to be generated for appropriate surveillance of genes of interest. This approach enables the effective identification of patients, disease groups and genomic variants that are amenable for functional assessment of mis-splicing through RNA-seq, and may help to improve the efficiency and accuracy of genomic diagnostic approaches.

The primary purpose of MRSD is to predict the likelihood of observing pathogenic splicing defects in a given gene and tissue, and we quantify the utility of three distinct biosamples in this manner for known monogenic disease genes (Figure 6). Through this analysis, we are able to highlight biosamples that may be most informative for RNA-seq based analysis datasets for specific disease subsets. Although our model is conservative (Figure 2), we demonstrate through MRSD-guided re-inspection of VUSs in ClinVar that it may be possible to use RNA-seq to clarify the effect of up to 2.4% of variants of uncertain significance (Figure 7a).

Other approaches to select genes amenable to functional analysis through RNA-seq include leveraging relative gene expression metrics (14, 20), or tools which assess the similarity of transcript isoforms between tissues, e.g. MAGIQ-CAT (7). We show that, whilst TPM values are well correlated with MRSD scores (Figure 3a-c), uneven sequencing coverage across the length of the transcript may, in some cases, falsely identify specific genes or splice junctions as being amenable to RNA-seq-based analysis (Supplementary Figure 5). 3’ sequencing bias, which is a known artefact of poly-A enriched mRNA sequencing (21-23), may elevate the risk of inaccurately selecting genes that could be surveyed through RNA-seq when considering TPM alone. Additionally, the normalization against sequencing depth that occurs during the calculation of TPM obscures information about raw read count, which is important when analyzing the utility of RNA-seq for clinical diagnostics. MRSD scoring, conversely, leverages variation in sample read depth to provide quantitative predictions about optimal sequencing depths.

On the other hand, the recently released tool MAGIQ-CAT (7) assesses the degree to which transcript isoforms in a sampled tissue accurately resemble those in the primary disease-affected tissue. However, MAGIQ-CAT primarily captures the degree of similarity between isoform structure and does not aim to provide a quantitative readout to guide the diagnostic route. Thus, a proxy tissue may be described as suitable for RNA-seq-based analysis despite having poor coverage of splice junctions. We envision that the use of both MAGIQ-CAT and MRSD could comprehensively capture information about the utility of RNA-seq, both in terms of similarity of isoform structure relative to the disease-affected tissue and in terms of the likelihood of observing disruptions to this structure.

There are several limitations of the current MRSD model, which could be incorporated into future work. Firstly, the MRSD model cannot directly be extended to predict the suitability of datasets to detect allele-specific expression biases and differential gene expression, which have been demonstrated to be evidence of pathogenic mechanisms in known disease-causing genes (10, 11, 14, 24). Although further investigations are required to quantify and prove this suitability, it is likely that genes with low MRSD scores (Figure 3d) are also amenable to investigations of differential gene expression and isoform imbalance.

Secondly, further extensions to the model could incorporate genomic background which influences gene expression profiles. For example, interferonopathies are a class of genomic immune disorders (25, 26) that are characterized by the aberrant upregulation of large numbers of transcripts belonging to so-called “interferon-stimulated genes” (25, 27). As a result of these wide-ranging impacts on their transcriptomes, MRSD predictions, which ostensibly represent the “normal” transcriptomic landscape, may not accurately reflect the degree of sequencing coverage for certain transcripts in patients with interferonopathies, or indeed other disease groups where disrupted expression of many transcripts is characteristic, such as disorders where chromatin structure (28, 29) or the function of the spliceosome (30-32) is disrupted. Moreover, the current MRSD model does not explicitly account for the presence of expression quantitative trait loci (eQTLs) or splicing quantitative trait loci (sQTLs) which are known to influence gene expression profiles (33-35). We have demonstrated that modulation in expression levels may disrupt our ability to reliably highlight pathogenic splicing events (Figure 5c). As a greater number of paired transcriptome and genomic datasets become available, we expect that MRSD scores can be generated in a dynamic manner to account for the presence of eQTLs, sQTLs or other modifiers of gene expression profiles.

Thirdly, our approach is built for a specific set of RNA-seq-based analyses; namely, the analysis of a selection of tissues by bulk short-read poly-A enrichment RNA-seq, followed by a specific bioinformatics analysis pipeline (13). This experimental RNA-seq approach currently remains widespread (3, 10, 13-15); however, our model may be readily applicable to RNA-seq generated using alternative methodologies, such as increased read length, with only minor variations in model performance (Supplementary Figure 3). As other technologies, such as long-read (36-38), single-cell (39, 40) and spatially resolved RNA-seq (41-44), become more prevalent in a clinical setting, appropriate control datasets must be generated to develop corresponding MRSD models. Similarly, recent research has shown noticeable improvements to diagnostic yield for neuromuscular disorders by conducting RNA-seq on *in vitro* myofibrils generated by a fibroblast-to-myofibril transdifferentiation protocol (45). Such patient-derived cell line approaches represent a promising avenue to scrutinize transcripts not otherwise observable in proxy tissues (31, 46). As these protocols gain wider use, generation of control RNA-seq data from healthy individuals using these approaches will be vital both to allow the generation of MRSD scores and to accurately assess pathogenicity of any identified mis-splicing events.

## Conclusions

In summary, the novel MRSD model presented here offers a gene-specific readout to predict the most suitable biosample for interrogation of splicing disruption at the transcript level. This may uncover previously unintuitive choices of biosample, as discussed above in the case of familial rhabdomyosarcoma (Figure 6c). The use of different biosamples is associated with different costs: while whole blood is routinely taken in the clinic, cell-based RNA-seq requires harvesting and culturing of patient cells, and muscle biopsy is an invasive procedure that is generally only undertaken if deemed necessary. Our tool may allow clinical staff to make informed decisions about the likely cost-benefit balance of RNA-seq analysis to ensure such costs are not incurred unnecessarily. We expect that the use of MRSD will allow effective and appropriate integration of RNA-seq into diagnostic genomic services, and ultimately improve variant interpretation and diagnostic yield.

## Methods

### Minimum required sequencing depth (MRSD) score

We generated a collated map of splice junction coverage for GTEx samples from three tissues (peripheral blood: *n* = 151; LCLs: *n* = 91; skeletal muscle: *n* = 184; see *RNA-seq data acquisition*, below), using established methods (Cummings et al., 2017). These samples were designated as *reference sets*. Our model considers the level of sequencing coverage for splice junctions in each tissue-specific reference set and calculates the minimum required sequencing depth (MRSD), in millions of uniquely mapping 75 bp reads, that would be required for the desired proportion of splice junctions in a given gene to be covered by a desired number of sequencing reads. Our model is dynamic, and can be adjusted by the user to account for customized levels of desired sequencing coverage per splicing junction, the proportion of splicing junctions covered, and the confidence level with which MRSD will generate datasets with the specified level of coverage (suggested usage of 95 or 99%).

MRSD is defined for a given gene in a given sample as:

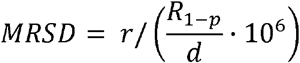

Where *r* is the desired level of read coverage across desired proportion *p* of splice junctions, *R* is the set of read counts supporting all junctions in the transcript of interest, and *d* is the total number of sequencing reads in the RNA-seq sample (by default, the number of uniquely mapping sequencing reads). The term *R*_1*-p*_ corresponds to the number of reads covering the junction with the “1 - *p*”-th-highest read count across all splice junctions in the transcript of interest.

MRSD scores have been generated for specified transcripts across all samples within the reference set in the three tissues of interest. The score at the X^th^ percentile position in the reference set list is returned as the MRSD, where X is termed the “confidence level” and is customizable by the user (default = 95%, Supplementary Methods 1).

### Transcript selection

MRSD can be calculated for any transcript sets of interest. Here, we utilized a hierarchy for transcript selection for all genes present in the GENCODE v19 human genome annotation (Supplementary Methods 2). We prioritized transcripts in the MANE v0.7 curated transcript list, providing that all splicing junctions were supported in the GENCODE v19 annotation. Genes without MANE transcripts were assigned composite transcripts, consisting of the union of all junctions found in transcripts in NCBI RefSeq transcripts. For genes that matched neither criteria, the union of all junctions present in all GENCODE v19-listed transcripts for that gene were used as the transcript model.

### Control RNA-seq data acquisition

FASTQs were downloaded from the Database of Genotypes and Phenotypes (dbGaP) under the project accessions phs000424.v8.p2 and phs000655.v3.p1.c1 for GTEx control individuals and neuromuscular disease patients, respectively. GTEx controls were selected for LCLs (*n* = 91), skeletal muscle (*n* = 184) and whole blood (*n* = 151) according to tissue-specific criteria (Supplementary Methods 3) to ensure use of only high-quality samples in generating control splicing datasets.

### In-house RNA-seq generation

RNA-seq datasets used to evaluate model performance were accessed from previously published datasets (13), under dbGaP project accession phs000655.v3.p1.c1, through international consortia (47), or for individuals in whom written informed consent was obtained and ethical approval for the study granted by Scotland A (refs: 06/MRE00/76 and 16/SS/0201), South Central-Hampshire A (ref: 17/SC/0026), South Central-Oxford B (ref:11/SC/0269) or South Manchester (ref: 11/H10003/3).

For in-house peripheral blood samples, RNA was extracted from PAXgene Blood RNA Kits and underwent poly-A enrichment library preparation using the TruSeq Stranded mRNA assay (Illumina) followed by 76 bp paired end sequencing using an Illumina HiSeq 4000 sequencing platform. For in-house LCL samples, RNA was extracted from pelleted LCLs thawed directly into TRIzol reagent (Invitrogen, 15596-026) using chloroform, and treated with TURBO DNase (Invitrogen, AM1907), both following the manufacturers’ instructions. RNA was prepared using the NEBNEXT Ultra II Directional RNA Library Prep kit (NEB #7760) with the Poly-A mRNA magnetic isolation module (NEB #E7490), according to manufacturer’s instructions, and 75bp paired end sequencing was performed using the Illumina NextSeq 550 sequencing platform. Ribosomal RNA depleted datasets were generated using RNA extracted via the PAXgene Blood RNA system, and 150bp paired end sequencing performed via Novogene (Hong Kong) using the NEBNext Globin and rRNA Depletion and NEBNext Ultra Directional RNA Library Prep Kits on a HiSeq 2000 instrument (Illumina). RNA samples from 20 LCLs were obtained from the kConFab consortium. Poly(A)-selected RNA was generated using the TruSeq Stranded mRNA Library Prep Kit (Illumina), and 150bp paired end reads created using the NextSeq 500 instrument (Illumina).

### Splice event identification

All FASTQs were aligned and processed as previously described (Cummings et al., 2017). Briefly, this analysis consisted of two-pass alignment using the STAR v2.4.2 aligner, marking of suspected PCR duplicates, and processing of the resultant alignments to generate tissue-by-tissue lists of splice junctions present within the cohort. Metrics for each splicing event were collected (Box 1), and splicing junctions were filtered to retain only those events that were unique to single samples (singletons) or that were present in multiple samples (non-singletons) but with an increased usage in the sample of interest, that is, with a higher normalized read count (NRC), than any control. The resulting list was ranked according to NRC fold change, with singletons with high read counts considered the most significant events. The resulting junctions were considered “events of interest”.

### Factors influencing the likelihood of aberrant splicing identification

To calculate how the level of background splicing aberrations was altered by sample size, each individual in the three control splicing datasets was processed using the above pipeline (13) and compared against 2000 bootstraps of 30, 60 and 90 controls each from their respective control tissue dataset with replacement. Events were then filtered to retain only those events for which the NRC was higher in the given individual than in any controls, and then counted for each bootstrap. Median counts for singleton and non-singleton events were then collated for each control group size. We selected 32 aberrant splicing events identified in neuromuscular patient RNA-seq data. From the genes in which we identified these variants, samtools was used to remove random subsets of reads in 10% intervals from each of these events to simulate variability in the number of reads generated for the gene of interest. The resulting datasets, exhibiting variable expression of a single gene, were then rerun through the splice analysis pipeline and the above metrics gathered for these simulated datasets.

### Genomics England PanelApp data collection

Tabulated versions of 284 gene panels were downloaded from the Genomics England PanelApp repository. Each panel was filtered to retain only genes assigned a “green” classification for that panel, representing the highest level of confidence of a real genotype-phenotype association.

### Curation of ClinVar variants of uncertain significance

A tabulated version of the comprehensive ClinVar variant listing (17) for January 2021 was downloaded and filtered to retain only those variants that were annotated as either “Uncertain significance” or “Conflicting interpretations of pathogenicity”. SpliceAI scores (v1.2.1; (18)) were generated for these variants and those with a score of 0.5 or greater retained for downstream analysis.

## Data Availability

The control datasets used to generate the MRSD model are available through the dbGaP repository as part of the GTEx v8 release (accession phs000424.v8.p2). Publicly available muscle-derived RNA-seq datasets to test the model are available at dbGaP (accession phs000655.v3.p1.c1). Source code will be made available upon publication. All MRSD scores are available at http://mcgm-mrsd.github.io/.

## Declarations

### Ethics approval and consent to participate

External datasets utilized in this study were accessed under dbGaP project accessions phs000655.v3.p1.c1 and phs000424.v8.p2. Informed written consent was obtained for all inhouse analyses, with ethical and study approval from South Central-Hampshire A (ref: 17/SC/0026), South Central-Oxford B (ref:11/SC/0269), South Manchester (ref:11/H10003/3) and Scotland A (refs: 06/MRE00/76 and 16/SS/0201) Research Ethics Committees.

### Consent for publication

No identifiable patient information is reported in this study.

### Authors’ contributions

The study was designed and coordinated by C.F.R., G.C.M.B., R.T.O, S.H., T.A.B. & J.M.E. All authors contributed genetic or phenotypic data. C.F.R. and J.E. wrote the manuscript. A.T. designed and implemented the MRSD web portal. All authors contributed to the editing and revision of the manuscript.

## Acknowledgements

We wish to thank Heather Thorne, Eveline Niedermayr, all the kConFab research nurses and staff, the heads and staff of the Family Cancer Clinics, and the Clinical Follow Up Study (which has received funding from the NHMRC, the National Breast Cancer Foundation, Cancer Australia and the National Institute of Health (USA)) for their contributions to this resource, and the many families who contribute to kConFab. kConFab is supported by a grant from the National Breast Cancer Foundation, and previously by the National Health and Medical Research Council (NHMRC), the Queensland Cancer Fund, the Cancer Councils of New South Wales, Victoria, Tasmania and South Australia, and the Cancer Foundation of Western Australia. We also wish to thank members of the Wessex Investigational Sciences Hub (WISH) Laboratory, Southampton, UK, for their help in facilitating RNA-seq of kConFab LCL samples (particularly Christopher Mattocks, Daniel Ward and Jade Forster), as well as the work of the University of Manchester Genomics Core Technology and Bioinformatics Facilities for their assistance in sample processing.

