## Supplementary Material for "MRSD: a novel quantitative approach for assessing suitability of RNA-seq in the clinical investigation of mis-splicing in Mendelian disease"


**Supplementary Table 1**
Summary of pathogenic splicing variants analyzed during this study 3

**Supplementary Table 2**
Summary of datasets used in this study 4

**Supplementary Figure 1**
Categories of potentially pathogenic splicing events and their representation in analytical pipeline output 5

**Supplementary Figure 2**
Sequencing depths of RNA-seq samples used for evaluation of MRSD model accuracy 6

**Supplementary Figure 3**
Effect of varying sequencing read length on MRSD model performance 7

**Supplementary Figure 4**
Evidence for 3' sequencing bias confounding the use of TPM as a guiding RNA-seq metric 8

**Supplementary Figure 5**
Exemplar events identified during pathogenic splice event analysis 9

**Supplementary Figure 6**
Relative gene expression level does not reflect the raw read coverage of transcript splice junctions 10

**Supplementary Figure 7**
Pairwise comparisons, by tissue, of MRSD scores for PanelApp disease genes 11

**Supplementary Figure 8**
Proportion of low-MRSD genes per tissue for all PanelApp panels, ordered by panel size

**a) Low-MRSD gene proportions for large panels (> 50 genes)** 12
**b) Low-MRSD gene proportions for medium panels (21-50 genes)** 13
**c) Low-MRSD gene proportions for small panels (11-20 genes)** 14
**d) Low-MRSD gene proportions for very small panels (≤ 10 genes)** 15

**Supplementary Figure 9**
Proportion of low-MRSD genes per tissue for all PanelApp panels, ordered alphabetically by panel name
- **a) Low-MRSD gene proportions for panels named A-E** 16
- **b) Low-MRSD gene proportions for panels named F-L** 17
- **c) Low-MRSD gene proportions for panels named M-X** 18

**Supplementary Figure 10**
The potential power of RNA-seq for resolving ClinVar variants of uncertain significance (VUSs) 19

**Supplementary Methods 1**
Illustration of MRSD calculation methodology 20

**Supplementary Methods 2**
Tiering methodology for selection of transcripts for MRSD generation 21

**Supplementary Methods 3**
Tissue-specific criteria for filtering of high-quality GTEx control RNA-seq datasets 22

**Supplementary Methods 4**
Sample IDs of GTEx samples used to generate control datasets
- **Skeletal muscle** 23
- **Whole blood** 25
- **EBV-transformed lymphocytes (LCLs)** 27
- **Cultured fibroblasts** 28

**References** 30

| Variant (HGVSg) | Gene | Source of RNA | Phenotype | TPM | MRSD (M reads) |
|---|---|---|---|---|---|
| chr2:152,355,017G>T | NEB | Skeletal muscle | Nemaline myopathy | 857.9 | 9.83 |
| chr2:152,389,953A>C | | | | | |
| chr2:152,544,805C>T | | | | | |
| chrX:31,790,694-31,798,498invdel | DMD | | Duchenne muscular dystrophy | 24.84 | 79.4 |
| chrX:32,274,692G>A | | | Myalgia, myoglobinuria | | |
| chr2:179,642,185G>A | TTN | | Multi/minicore congenital myopathy | 349.5 | 47.63 |
| chr21:47,409,881C>T | COL6A1 | | Collagen VI-related dystrophy | 56.02 | 16.25 |
| chr21:47,409,881C>T | | | | | |
| chr19:38,958,362C>T | RYR1 | | Congenital fiber-type disproportion | 425.5 | 3.45 |
| chr1:46,655,129C>A | POMGNT1 | | α-Dystroglyanopathy | 29.26 | 6.01 |
| chr17:41,199,655C>G | BRCA1 | LCL | Inherited breast cancer susceptibility | 19.985 | 217.19 |
| chr17:41,246,879T>C | | | | | |
| chr17:41,246,879T>C | | | | | |
| chr17:41,246,879T>C | | | | | |
| chr17:41,258,551C>A | | | | | |
| chr13:32,945,238G>A | BRCA2 | | | 10.16 | Unfeasible |
| chr13:32,969,074A>T | | | | | |
| chr19:33,892,776C>T | PEPD | Whole blood | Prolidase deficiency | 18.89 | 28.31 |
| chr20:35,526,363C>G | SAMHD1 | | Aicardi-Goutières syndrome | 48.53 | 24.68 |
| chr23:153,997,595G>A | DKC1 | | Dyskeratosis congenita | 102.01 | 8.59 |

**Supplementary Table 1.** *Summary of pathogenic splicing events analyzed in this study.* All co-ordinates are given in relation to the GRCh37 genome build. TPM, transcripts per million; MRSD, minimum required sequencing depth.

| Tissue | No. samples | Source | Sequencing type | Usage |
|---|---|---|---|---|
| Blood | 151 | GTEx | 75-bp paired end poly-A enrichment, Illumina | Generation of MRSD model, bootstrapping analysis of event counts |
| LCL | 91 | | | |
| Muscle | 184 | | | |
| Blood | 1 | Inhouse | 150-bp paired end globin depletion, Illumina | Collation of known pathogenic mis-splicing events |
| | 12 | | 75-bp paired end poly-A enrichment, Illumina | Collation of known pathogenic mis-splicing events & MRSD model validation |
| LCL | 20 | | 150-bp paired end poly-A enrichment, Illumina | Collation of known pathogenic mis-splicing events |
| | 4 | Inhouse | 75-bp paired end poly-A enrichment, Illumina | MRSD model validation |
| Muscle | 52 | Previously published data [1] | 75-bp paired end poly-A enrichment, Illumina | Collation of known pathogenic mis-splicing events, downsampling of pathogenic events & MRSD model validation |

**Supplementary Table 2.** *Summary of RNA-seq datasets utilized in this study*. RNA-seq datasets derived using different methodologies were used for various aspects of this study. All data used to generate the MRSD model was based on data from the GTEx consortium across all three analyzed tissues.

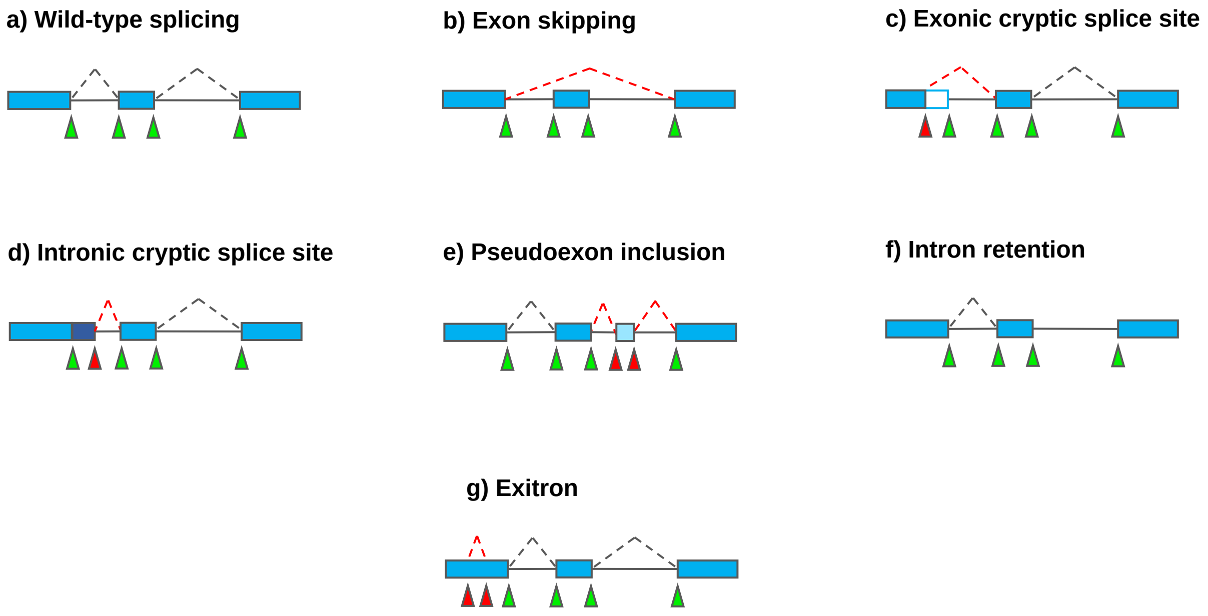


**Supplementary Figure 1.** *Categories of potentially pathogenic splicing events and their representation in analytical pipeline output.* Disruption of (**a**) wild-type splicing may lead to (**b**) skipping of one or more exons, the creation of novel splice sites in (**c**) exonic or (**d**) intronic regions that may outcompete the canonical sites, or result in (**e**) the generation of an intronic pseudoexon. (**f**) Splicing may be abrogated completely, leading to total retention of the intron. (**g**) Within longer exons, creation of a novel splice site may lead to a so-called “exitron”, whereby a central portion of the exon is absent from the final transcript. Green triangles indicate canonical splice sites; red triangles indicate non-canonical sites.

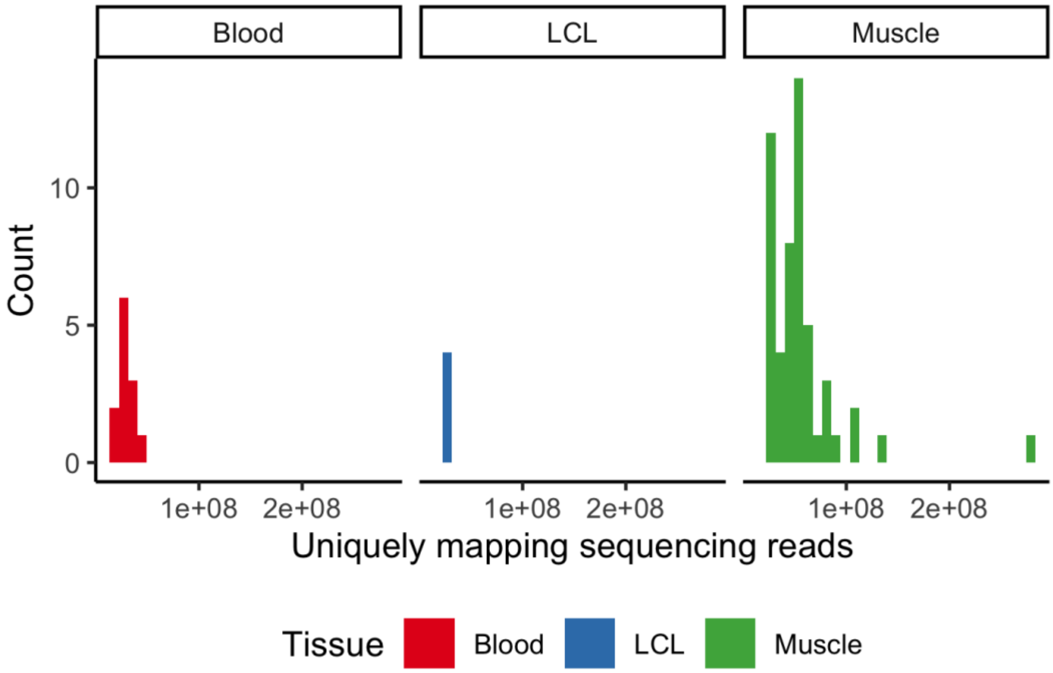


**Supplementary Figure 2.** *Sequencing depths of RNA-seq samples used for evaluation of MRSD model accuracy.* Whole blood (*n* = 12), LCL (*n* = 4) and skeletal muscle (*n* = 52) RNA-seq samples were derived from in-house or previously published data [1] for validation of the MRSD model efficacy. Sequencing depths across the three tissues ranged from 20.6-281.5 M uniquely mapping reads.

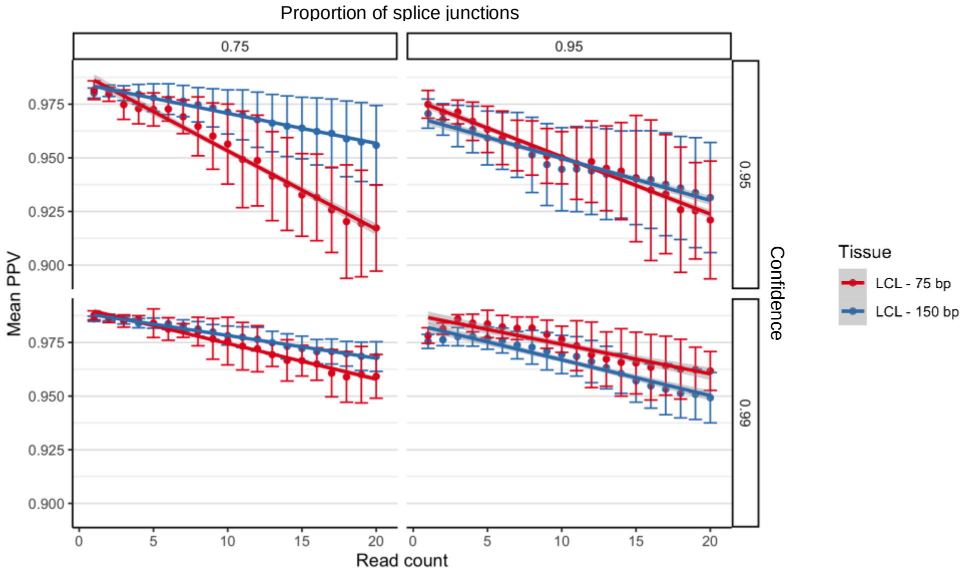


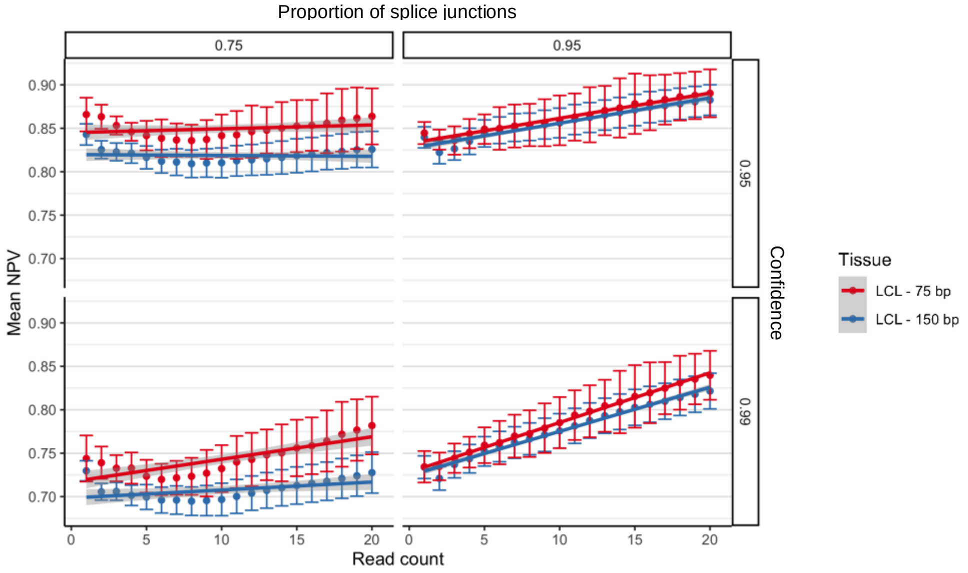


**Supplementary Figure 3.** ***Effect of varying sequencing read length on MRSD model performance.*** Despite being derived from 75 bp paired end RNA-seq data, MRSD scores show similar performance when applied to 75 or 150 bp paired end read-based RNA-seq, both in terms of (top) PPV and (bottom) NPV. When specifying 75% splice junction coverage, MRSD PPV is generally higher when the model is applied to 150 bp read-based data. This likely reflects the fact that junctions predicted to be sufficiently covered by 75 bp reads will be more likely to be sufficiently covered by reads of greater length, and so positive predictions are more likely to hold true when applied to longer-read data. We also observe that NPV for 150 bp read datasets is lower than that for 75 bp across all 4 parameter combinations; conversely to PPV, this is possibly because transcripts not sufficiently covered by 75 bp reads are more likely to be sufficiently covered by 150 bp reads, thus making negative predictions less likely to hold true in longer-read data. In most cases, differences in model performance between 75 and 150 bp is low, suggesting MRSD may, in some cases, provide a suitable approximation of transcript coverage in RNA-seq datasets with read lengths different to those used to construct the model.

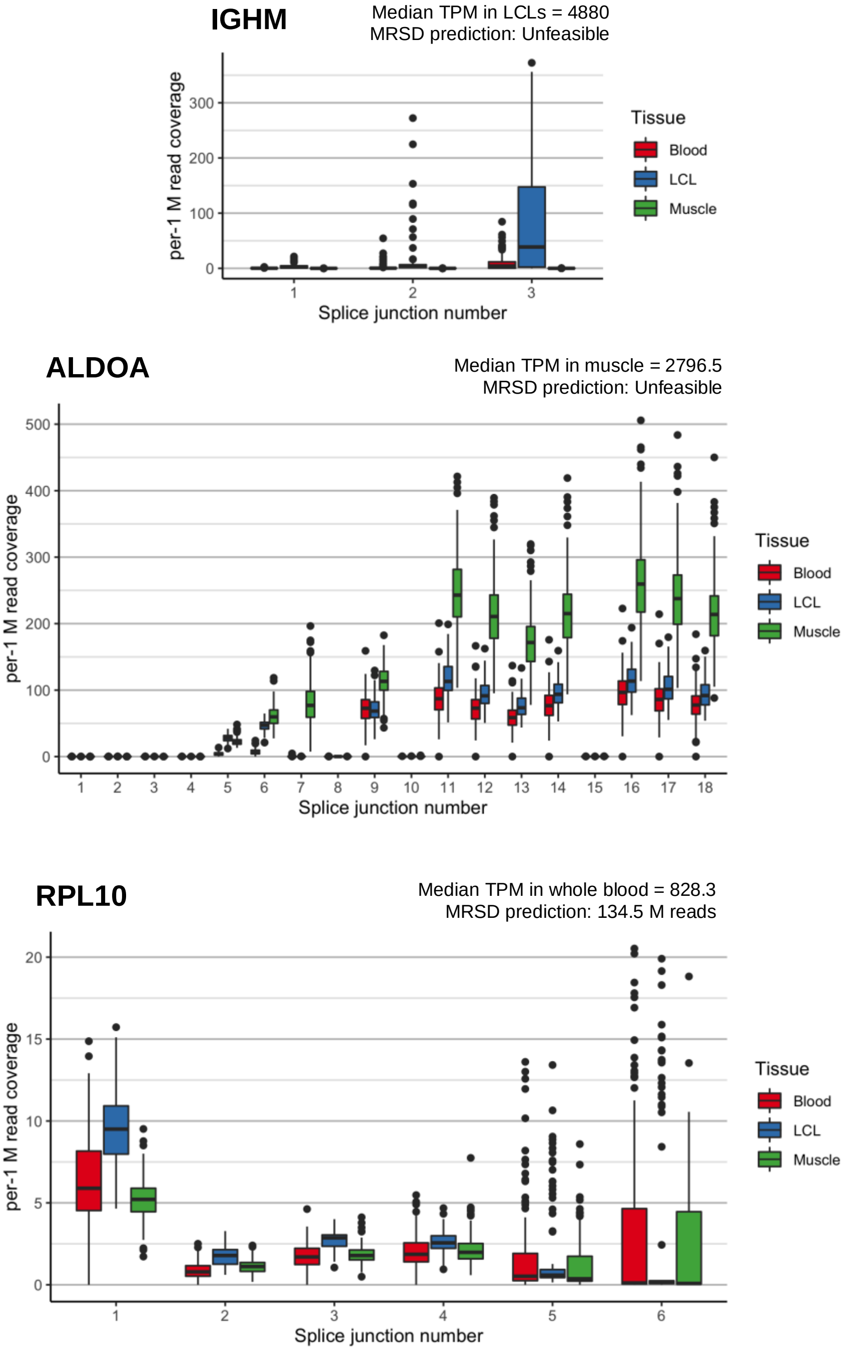


**Supplementary Figure 4.** *Evidence for 3' sequence bias confounding the use of TPM as a guiding RNA-seq metric.* Analysing the number of reads (per 1 M uniquely mapping input reads) mapping to individual splice junctions within three genes with substantial TPM-MRSD discrepancy demonstrates that highly expressed genes may exhibit biased coverage of splice junctions. For IGHM (top) and ALDOA (middle) in LCLs and muscle, respectively, a sufficient proportion of junctions towards the 3' end of the transcript have no read support in a sufficient number of patients, resulting in an MRSD prediction of "unfeasible", despite high coverage of other junctions within the same transcript. Coverage of the final two splice junctions in RPL10 (bottom) in LCL-based RNA-seq data is low but not non-zero in many patients, giving a feasible but high MRSD prediction. In some cases, this bias may result from artefacts of library preparation, or may possible reflect genuine isoform shifts in the given tissue. Higher splice junction numbers represent junctions closer to the 3' end of transcripts.

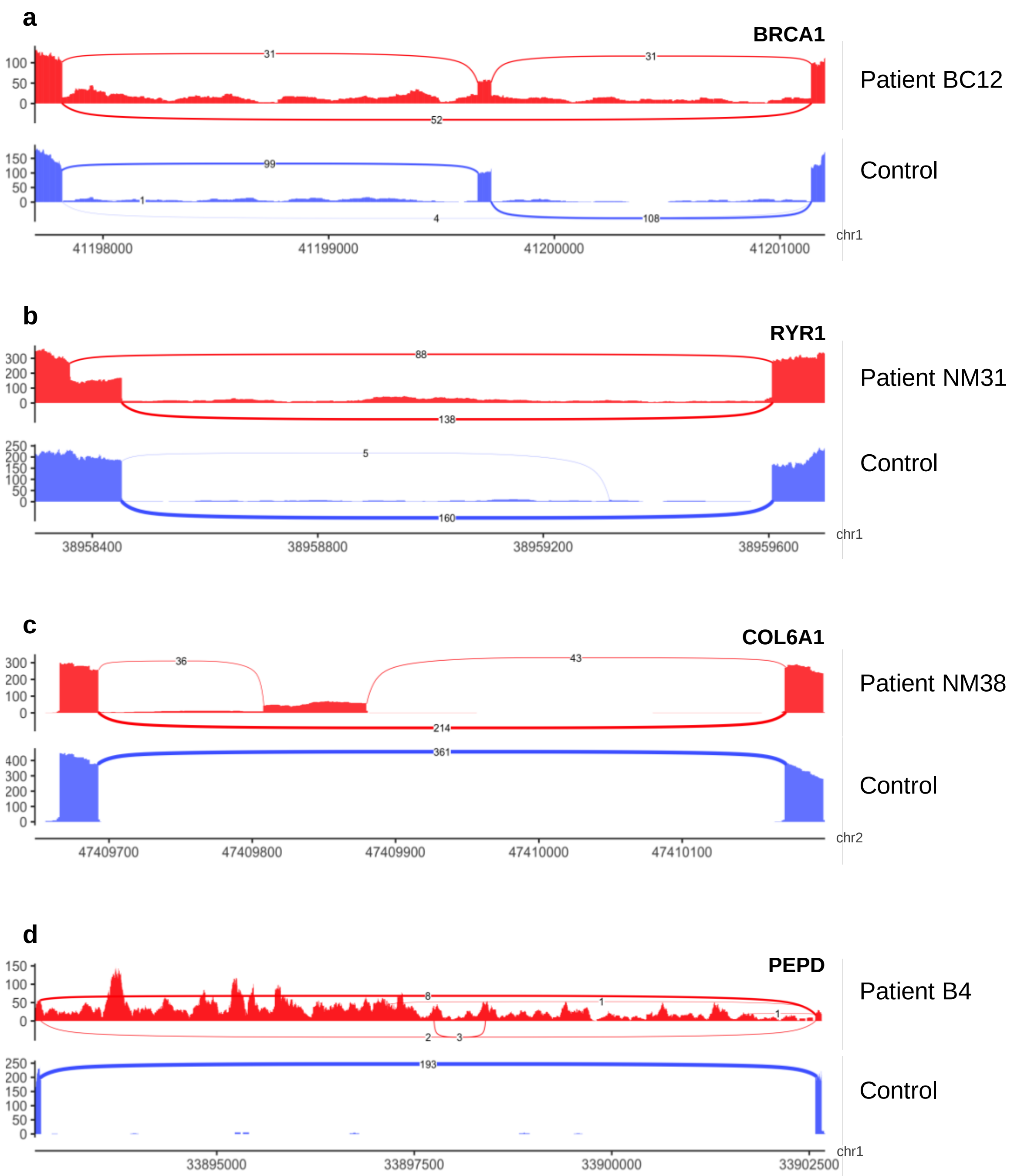


**Supplementary Figure 5.** *Exemplar events identified during pathogenic splice event analysis.* Selected Sashimi plots for (**a**) exon skipping, (**b**) exonic splice gain, (**c**) pseudoexonization and (**d**) intron retention events identified as the cause of disease in our patient datasets. The presence of aberrant splice junctions with outlying event metrics allowed flagging of these as potentially pathogenic. For (**d**), the intron retention event was identified from the 2 reads supporting usage of an extremely weak alternative splice acceptor four bases downstream of the abrogated canonical acceptor; however, in the absence of any aberrant splicing events, intron retention events are more difficult to identify from RNA-seq data using current bioinformatics pipelines.

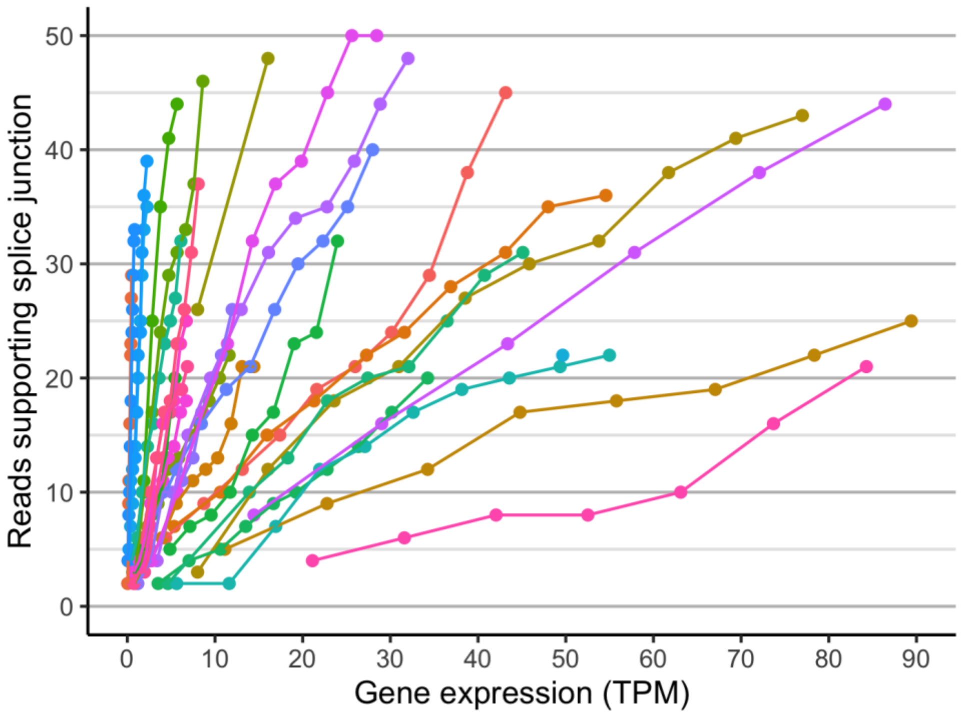


**Supplementary Figure 6.** *Relative gene expression level does not reflect the raw read coverage of transcript splice junctions.* When simulating decreased gene expression by downsampling reads in genes containing novel splicing events identified in upstream analysis, it emerged that expression of a gene (in transcripts per million, TPM) does not directly correlate with the number of reads supporting splice junctions in that gene. Among the events supported by 8 reads, for example, gene expression ranged from 0.17-52 TPM. This may be accounted for by variation in the proportion of transcripts containing the event, variation in the coverage across the length of a transcript (as shown in Supplementary Figure 4), or variation in the depth to which a sample has been sequenced. Thus, when specifying a metric threshold above which we expect splice aberration to be observable, relative expression level may not appropriately represent expected read support. Axes are limited for ease of visualization.

**Supplementary Figure 7.** *Pairwise comparisons, by tissue, of predicted MRSD scores for PanelApp disease genes.*

### a) MRSD predictions in muscle vs. blood

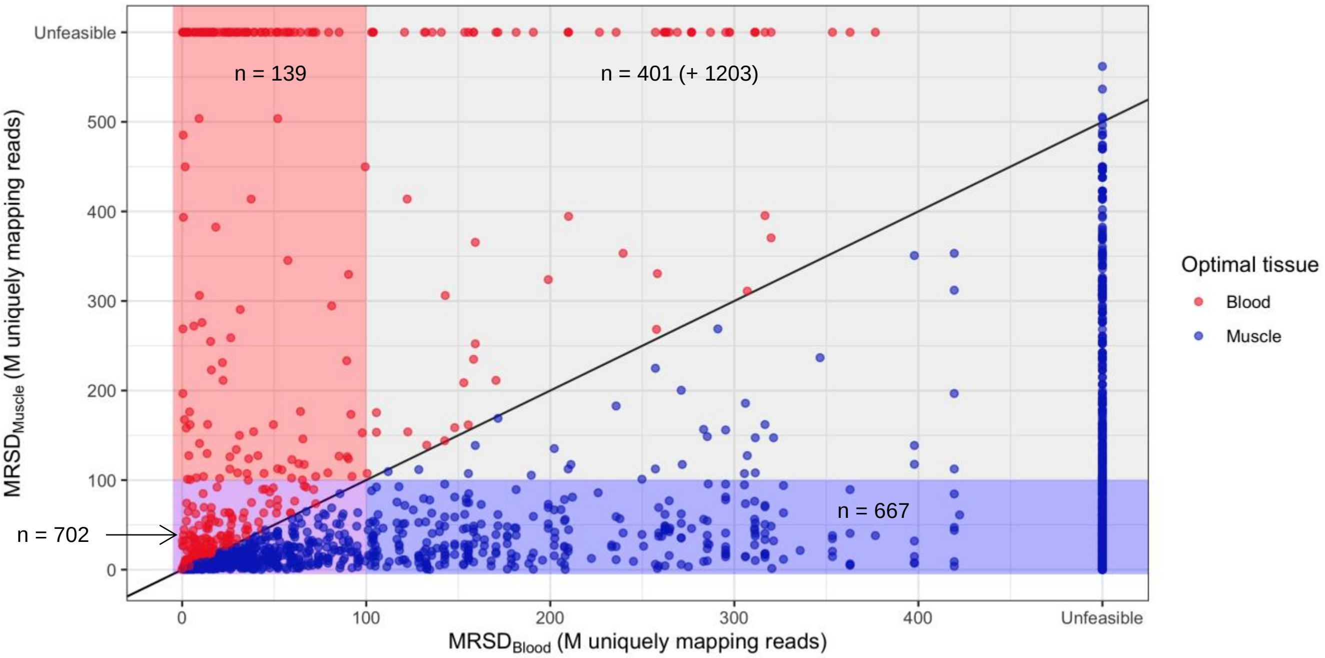


### b) MRSD predictions in LCLs vs. muscle

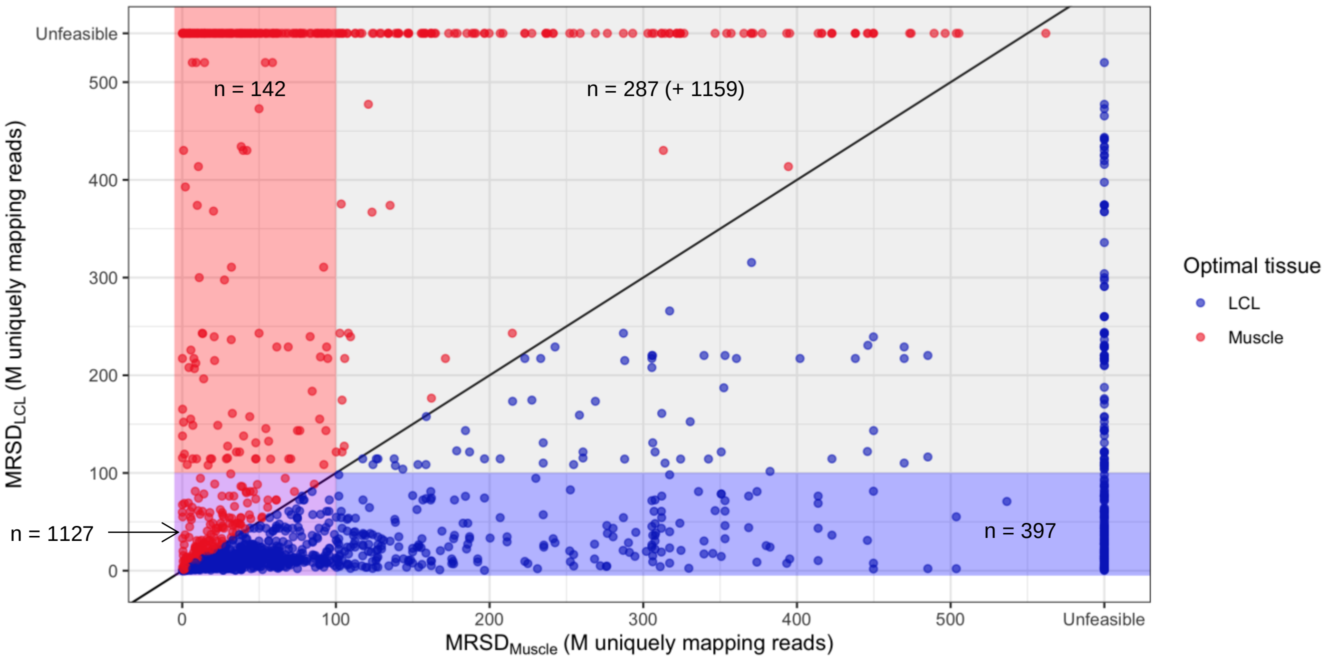

**Supplementary Figure 8.** *Proportion of low-MRSD genes per tissue for all PanelApp panels, ordered by panel size.*

### a) Low-MRSD gene proportions for large panels (> 50 genes)

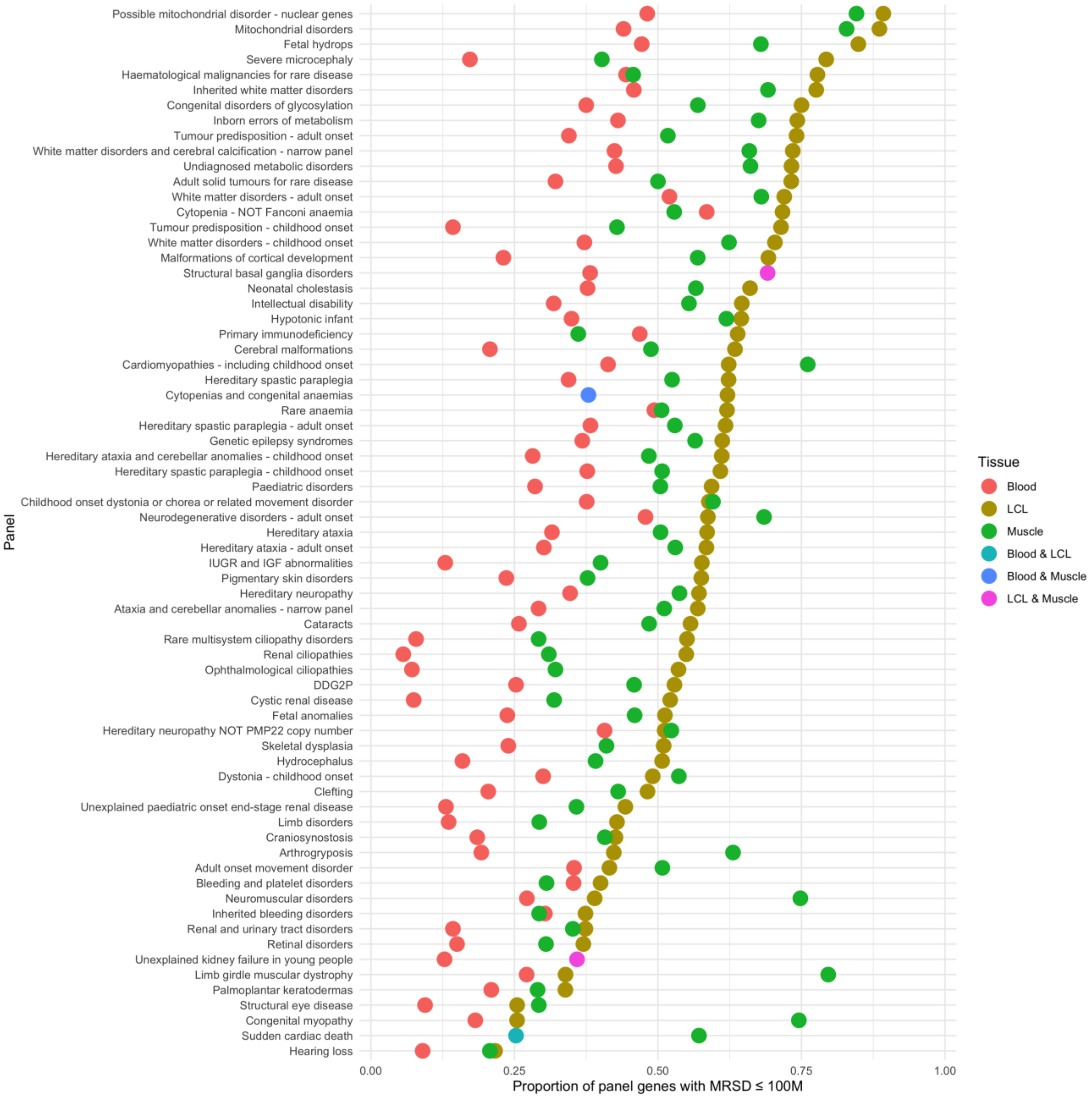

**b) Low-MRSD gene proportions for medium panels (21-50 genes)**

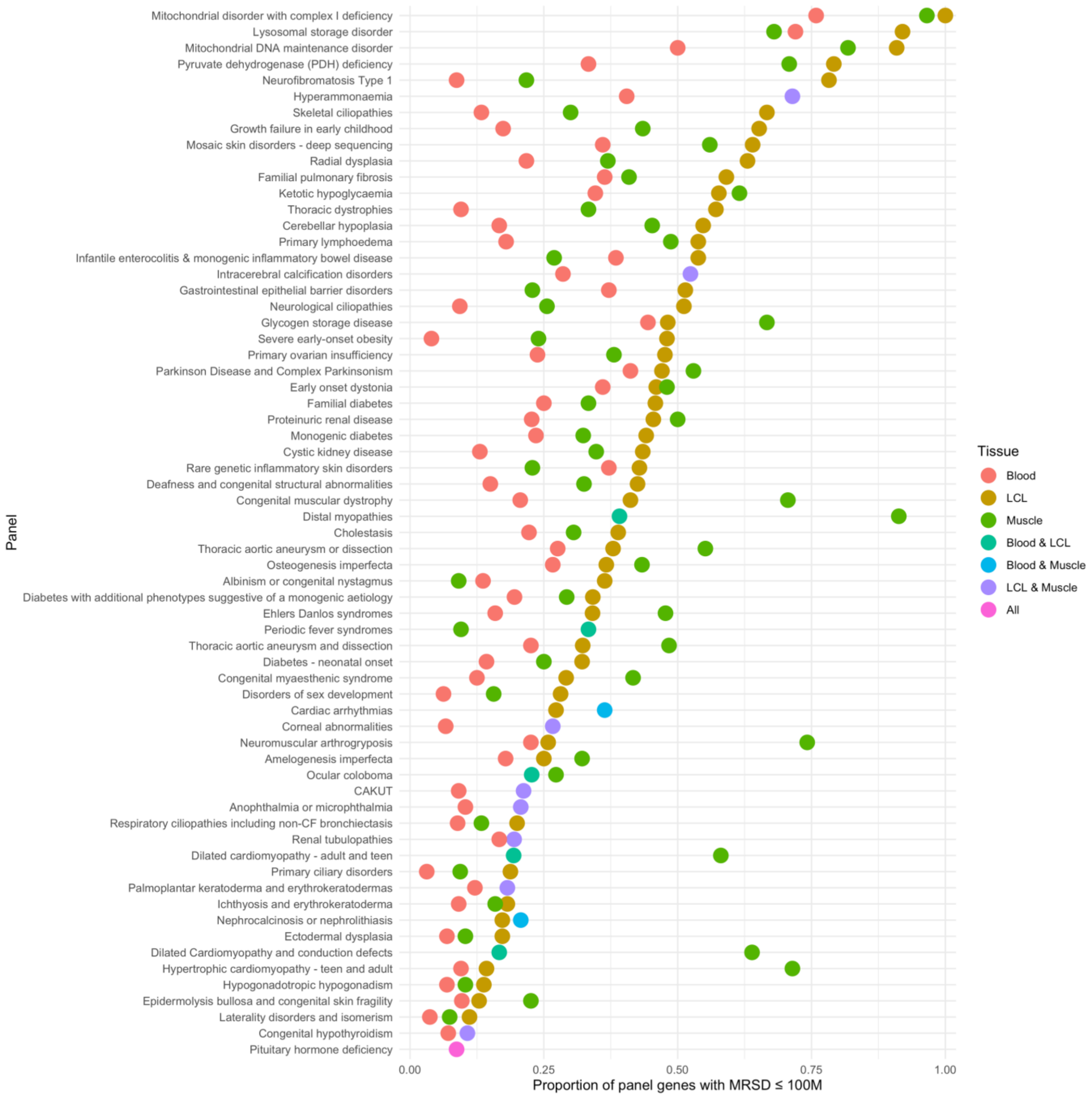

### c) Low-MRSD proportions for small panels (11-20 genes)

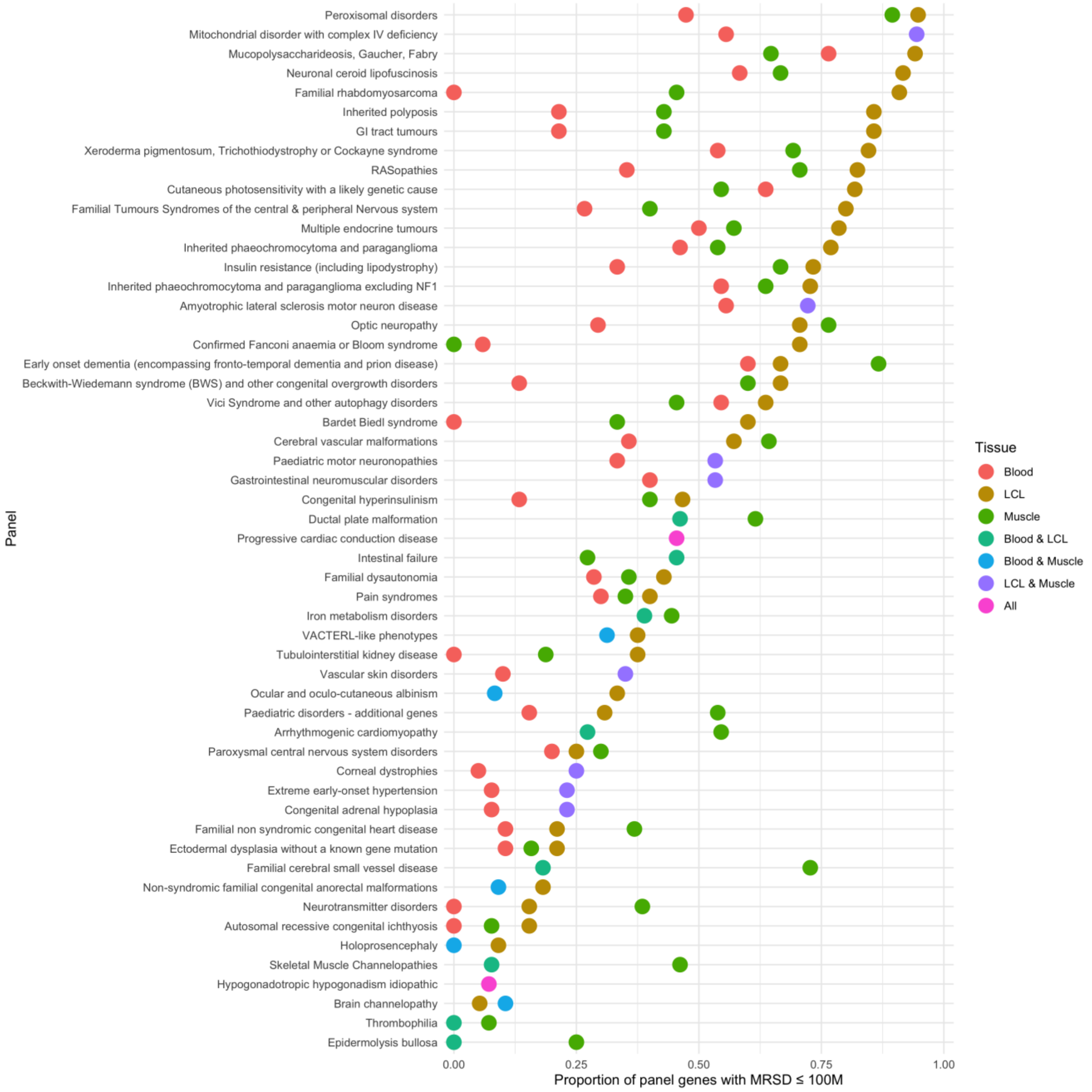

### d) Low-MRSD gene proportions for very small panels (≤ 10 genes)

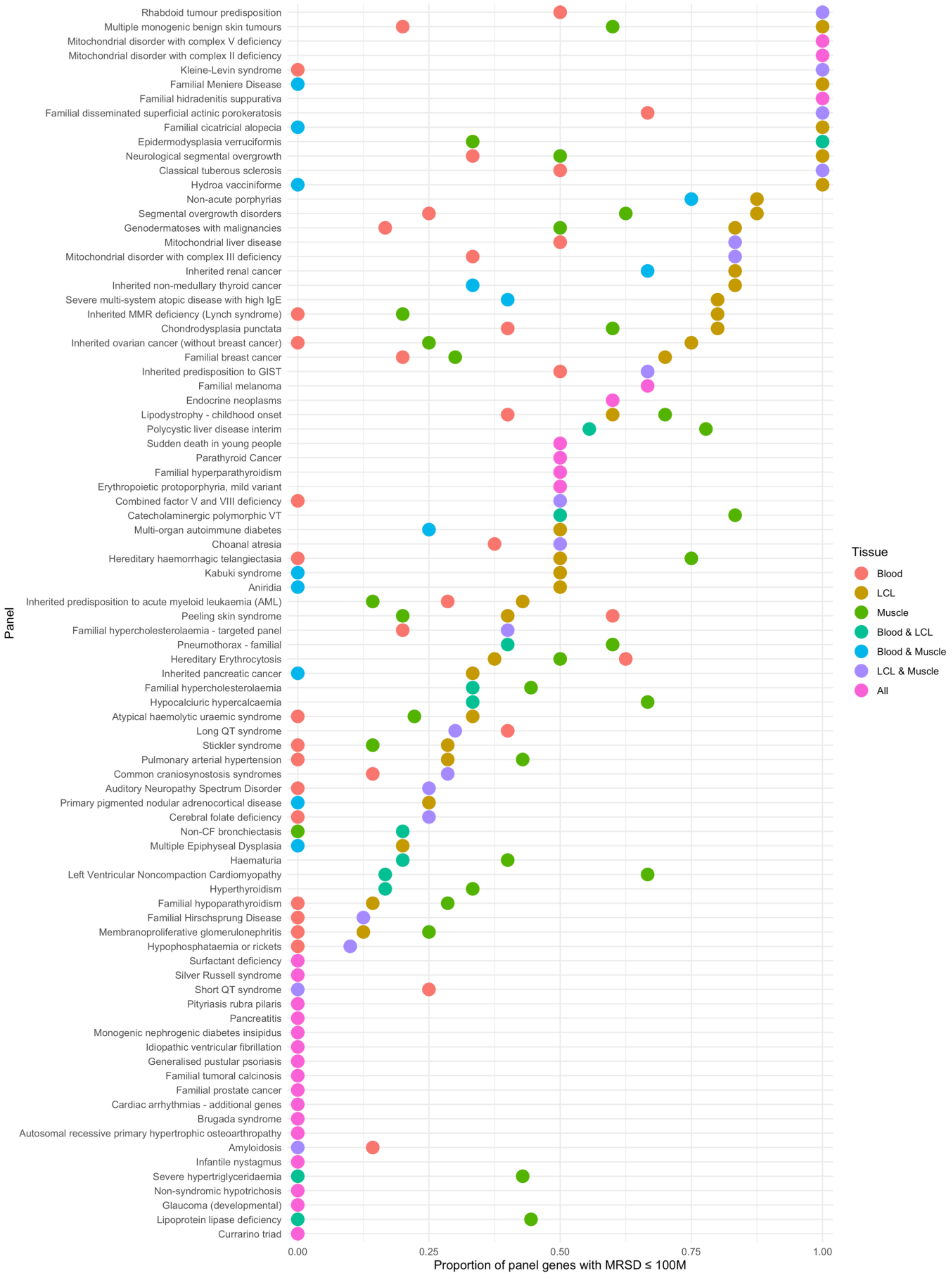

**Supplementary Figure 9.** *Proportion of low-MRSD genes per tissue for all PanelApp panels, ordered alphabetically by panel name.*

#### a) Low-MRSD gene proportions for panels named A-E

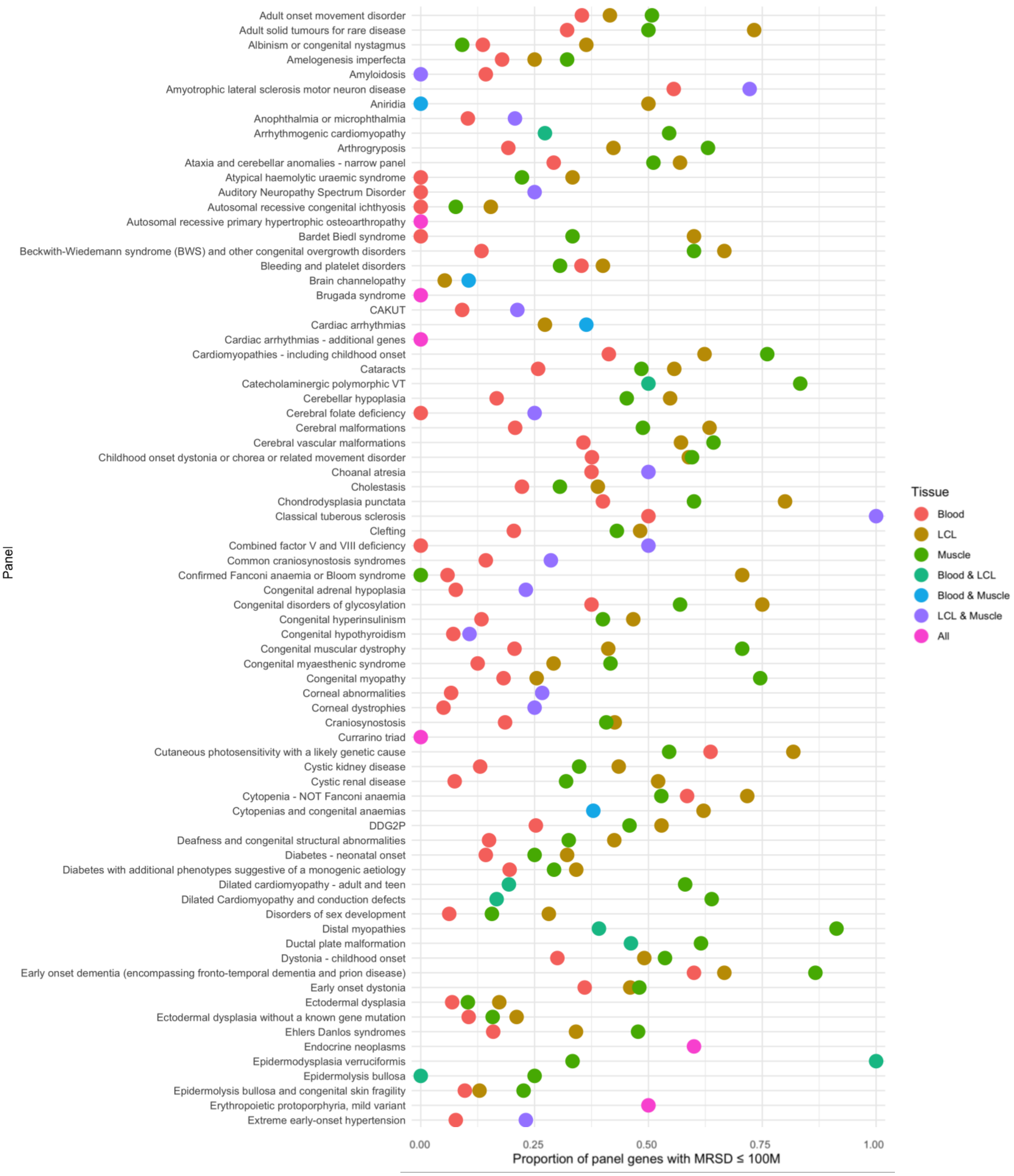

### b) Low-MRSD proportions for panels named F-L

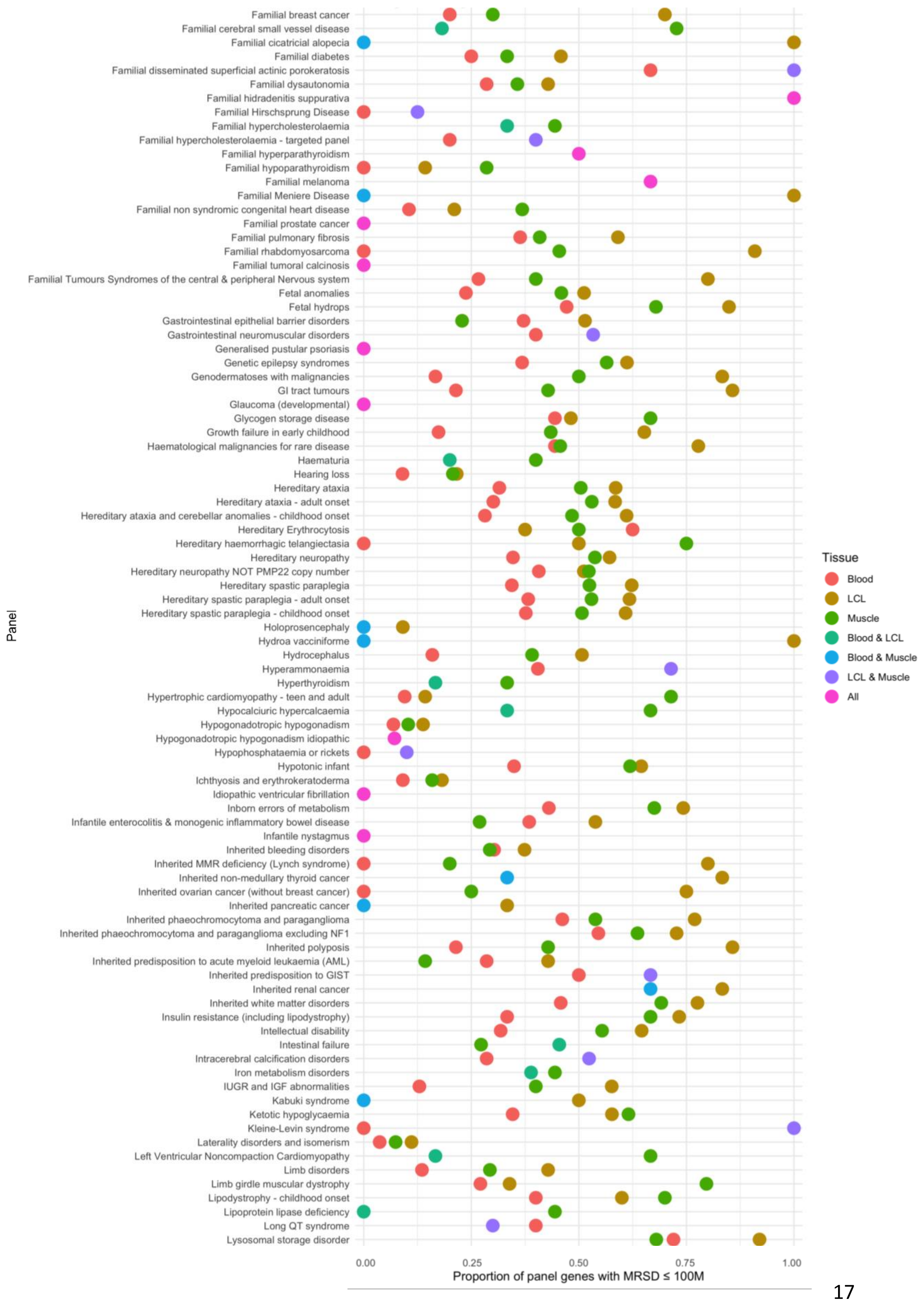

### c) Low-MRSD gene proportions for panels named M-X

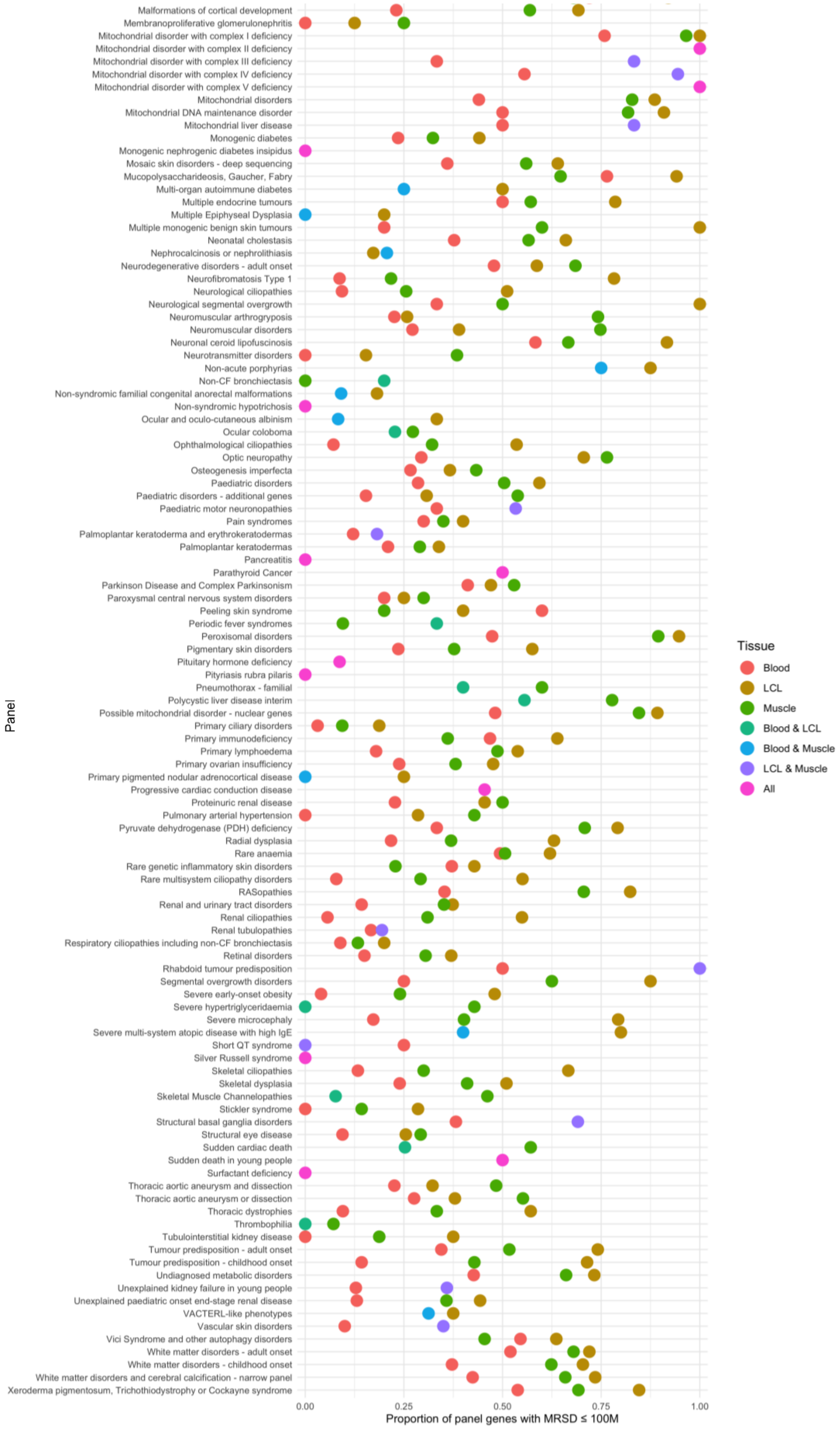

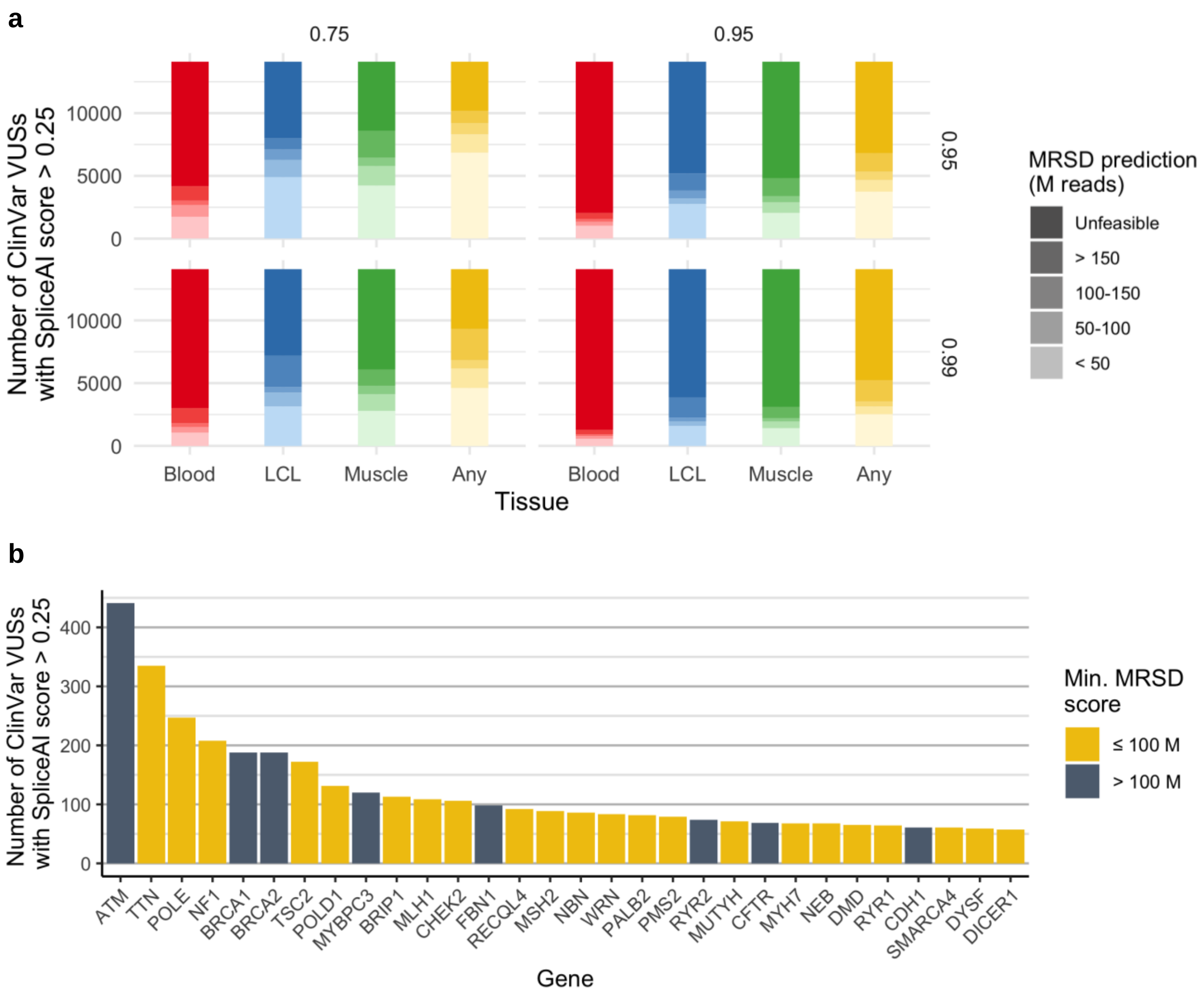


**Supplementary Figure 10.** *The potential power of RNA-seq for resolving ClinVar variants of uncertain significance (VUSs).* Similarly to Figure 7 (main text), we analyzed the distribution of MRSD predictions for genes harbouring ClinVar VUSs with SpliceAI predictions > 0.25 [2]. Of the 185,119 ClinVar variants annotated as "Uncertain significance" or "Conflicting interpretations of pathogenicity", 13,602 (7.3%) passed this threshold, and so were deemed "predicted splice-impacting". (**a**) Across all parameter combinations, a greater number of variants were in genes that were predicted low-MRSD (MRSD ≤ 100 M reads) in LCLs than in any other tissue. Dependent on parameter stringency, between 22.3% and 59.2% of variants were in genes predicted low-MRSD in at lest one of the three tissues. (**b**) Among the 30 genes containing the largest number of predicted splice-impacting VUSs in ClinVar, 22 were predicted low-MRSD in at least one of the three tissues.

**Supplementary Methods 1.** *Illustration of MRSD calculation methodology.* MRSD scores utilize the level of read coverage supporting the existence of splice junctions in control RNA-seq datasets to predict the depth of sequencing required to achieve a specified level of splice junction coverage. In a given individual and gene:

1. Read coverage values are collated across all splice junctions in the transcript model for that gene (see Supplementary Methods 2, below)
2. Each of these values is divided by the sequencing depth – by default defined as the number of uniquely mapping sequencing reads – and multiplied by $10^6$ to produce a per-1 M read coverage value for each junction
3. The desired level of read coverage is divided by the per-1 M read coverage value of the splice junction with the *X*'th percentile lowest read coverage, which gives the depth of sequencing that would be required for X% of junctions to be covered with the desired number of reads or higher. This figure is the sample-specific MRSD.

This process is iterated across all control RNA-seq samples to generate a list of sample-specific MRSDs, and a global MRSD is then derived by taking the *Z*'th percentile highest prediction from among these (*Z* corresponding to the so-called *confidence level*, i.e. the proportion of control RNA-seq samples for which that level of sequencing would have sufficiently covered that gene).

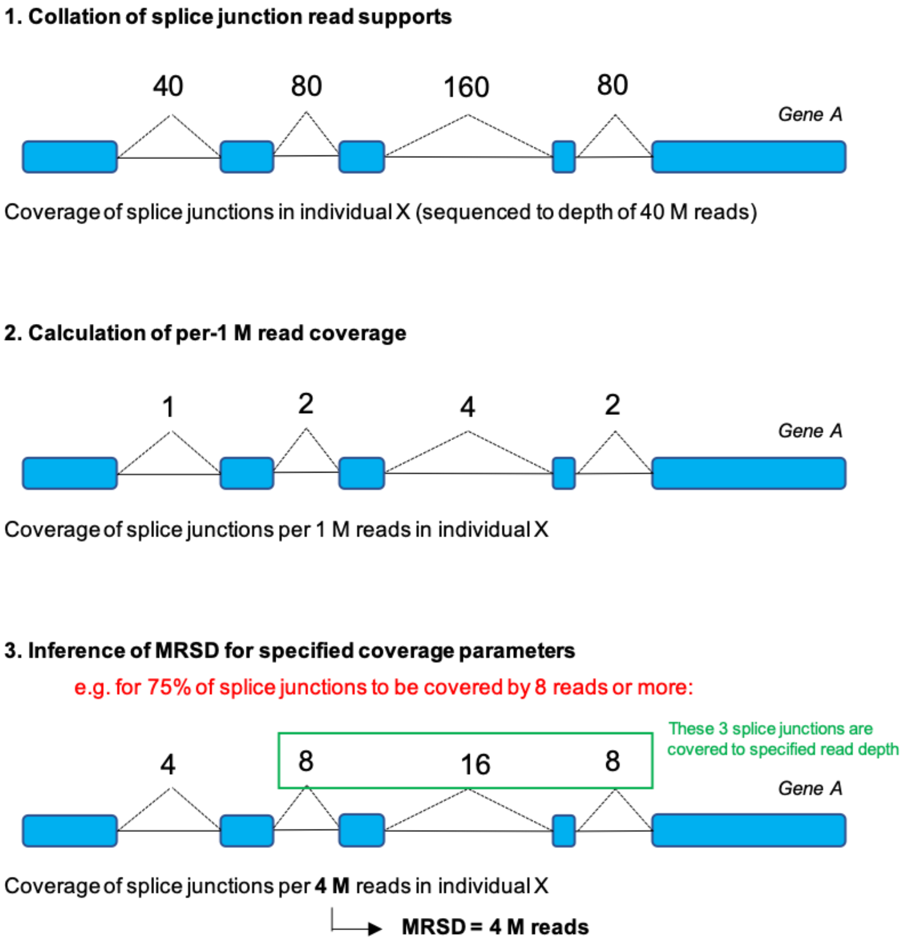

**Supplementary Methods 2.** *Tiering methodology for selection of transcripts for MRSD generation.* To calculate MRSD values for all protein-coding genes, a single transcript model was established for each gene. Firstly, transcripts present in the MANE v0.7 curated transcript set were selected for genes where these existed, provided the co-ordinates of all splice junctions in that transcript (given in relation to the GRCh38 reference genome) mapped back to known junctions in build GRCh37. For genes where these conditions were not met, transcript models were formed from the union of all junctions present in all RefSeq transcripts listed for that gene on Ensembl BioMart. Finally, for any genes lacking a corresponding RefSeq transcript, a transcript model was derived consisting of the union of all junctions present in all transcripts assigned to that gene in the GENCODE v19 annotation.

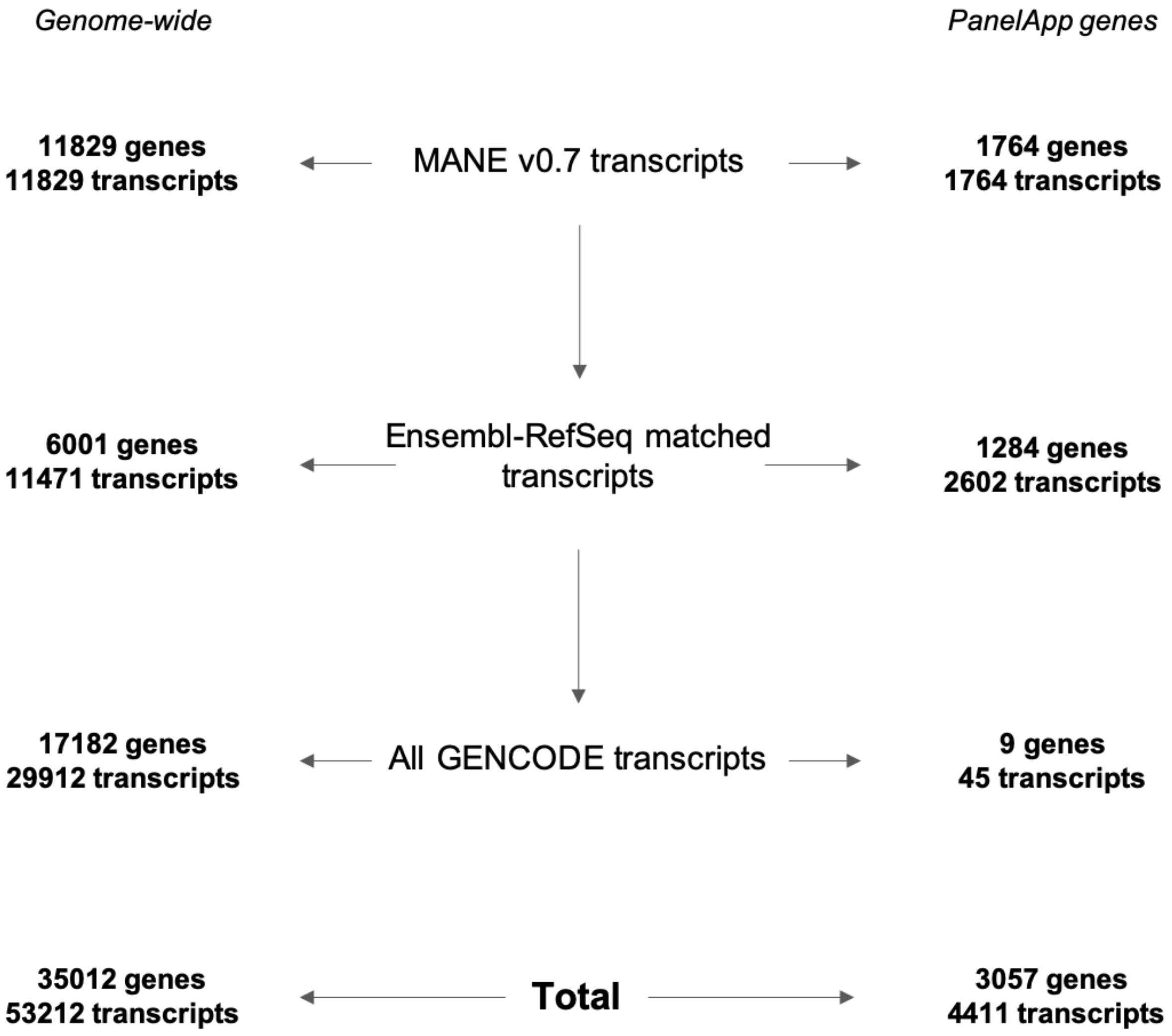

**Supplementary Methods 3.** *Tissue-specific criteria for filtering of high-quality GTEx control RNA-seq datasets.* Filtering of GTEx controls was conducted to select the highest quality samples based on the below tissue-specific parameters. Parameters were selected and adjusted on a tissue-by-tissue basis to exclude metric outliers and samples that may confound analysis of pathogenic splicing events (e.g. excluding cancer patients from LCL control cohorts, in which inherited breast cancer was studied). The corresponding column names in the GTEx v8 sample attribute (pht002743.v8) and subject phenotype (pht002742.v8) files are italicized.

*Skeletal muscle (as listed in [1])*

- RNA integrity number/RIN (*SMRIN*): between 6-9
- Sample ischemic time (*SMTSISCH*): <720 (i.e. <12 hours)
- Hardy scale (*DTHHRDY*): 0, 1 or 2, corresponding to sudden deaths
- Age (*AGE*): <50
    - Unless BMI <30

*Whole blood*

- Samples included in GTEx analysis freeze, corresponding to higher quality samples (*SMAFRZE*): not flagged EXCLUDE due to technical issues
- RIN (*SMRIN*): between 6-9
- Sample ischemic time (*SMTSISCH*): <0
- Hardy scale (*DTHHRDY*): 0, 1 or 2

*EBV-transformed lymphocytes (LCLs)*

- *SMAFRZE*: not flagged EXCLUDE due to technical issues
- RIN (*SMRIN*): > 9
- *MHCANCER5*, *MHCANCERC* and *MHCANCERNM* all 0 to eliminate all non-metastatic cancers and all cancers in the past 5 years or current
- *DTHHRDY*: 0, 1 or 2
- No reported history (*MHGENCMT*) of:
    - Breast cancer
    - Ovarian cancer
    - Pancreatic cancer
    - Prostate cancer
    - Colorectal cancer
    - No patients filtered out through this criterion

*Cultured fibroblasts*

- As for EBV-transformed lymphocytes, except with the addition of the following:
    - RIN (*SMRIN*) > 9.7
    - Uniquely mapping reads (*MPPDUN*): > 60 M

**Supplementary Methods 4.** *Sample IDs of GTEx samples used to generate control datasets*.

**Skeletal muscle** *(as listed in [1])*

GTEX-111CU-2026
GTEX-111YS-2326
GTEX-1122O-2426
GTEX-113JC-2726
GTEX-117YX-2526
GTEX-11DXX-2726
GTEX-11DXZ-2426
GTEX-11EM3-2126
GTEX-11EMC-2626
GTEX-11EQ9-2126
GTEX-11I78-2426
GTEX-11LCK-1226
GTEX-11NSD-2026
GTEX-11P81-2526
GTEX-11P82-1826
GTEX-11VI4-1926
GTEX-11WQC-2626
GTEX-11WQK-0726
GTEX-11XUK-2226
GTEX-11ZTT-2626
GTEX-11ZVC-2726
GTEX-1211K-2126
GTEX-12BJ1-2526
GTEX-12C56-1926
GTEX-12WSJ-1726
GTEX-12WSN-2526
GTEX-12ZZX-0326
GTEX-12ZZY-0626
GTEX-13111-2226
GTEX-1314G-1726
GTEX-131XF-2326
GTEX-131XG-2326
GTEX-132AR-1026
GTEX-132NY-0726
GTEX-1339X-2426
GTEX-133LE-2026
GTEX-1399Q-2426
GTEX-1399R-2526
GTEX-1399S-2726
GTEX-1399U-2526
GTEX-139D8-0726
GTEX-139UW-2626
GTEX-139YR-2526
GTEX-13CF3-1826
GTEX-13D11-2526
GTEX-13FH7-2126
GTEX-13FHO-0726
GTEX-13FTW-2326
GTEX-13FTY-0226
GTEX-OIZH-1626
GTEX-OOBJ-1626
GTEX-P4PP-1626
GTEX-P4PQ-1626
GTEX-P78B-1626
GTEX-POMQ-1926-SM-3NB1Y
GTEX-POYW-0526-SM-2XCEY
GTEX-PSDG-0426
GTEX-PWCY-2026
GTEX-Q2AH-1826-SM-2S1Q2
GTEX-Q734-2026-SM-3GADA
GTEX-QCQG-2126-SM-2S1P8
GTEX-QDVN-2426-SM-2S1Q4
GTEX-QV44-2026-SM-2S1RD
GTEX-R53T-1826-SM-3GIJX
GTEX-R55D-0626-SM-3GAD5
GTEX-S32W-2326-SM-2XCAW
GTEX-S33H-2226
GTEX-S7SF-2026-SM-3K2AS
GTEX-SNMC-1426-SM-2XCFM
GTEX-SUCS-1626-SM-32PLS
GTEX-T5JC-0626-SM-3NMA6
GTEX-T5JW-1826-SM-3GAE1
GTEX-TKQ2-0826-SM-33HB6
GTEX-TML8-1826-SM-32QOR
GTEX-TMMY-0426-SM-33HBB
GTEX-U3ZG-0326-SM-47JXN
GTEX-U3ZH-1926-SM-4DXTR
GTEX-U3ZM-1226-SM-3DB9G
GTEX-U4B1-1626-SM-3DB8N
GTEX-UJHI-1726-SM-3DB9B
GTEX-UJMC-1826-SM-3GADT
GTEX-VUSG-2626-SM-4KKZI
GTEX-WHPG-2226-SM-3NMBO
GTEX-WHSB-1826-SM-3TW8M
GTEX-WOFM-1326-SM-3MJFR
GTEX-WRHK-1626-SM-3MJFH
GTEX-WRHU-0826-SM-3MJFN
GTEX-WXYG-2526-SM-3NB3F
GTEX-WY7C-2526-SM-3NB2N
GTEX-WZTO-0826-SM-3NM8Q
GTEX-X4XY-0626-SM-4E3IN
GTEX-X638-0326-SM-47JY1
GTEX-X88G-0326-SM-47JZ4
GTEX-XBEC-0626
GTEX-XBED-2626-SM-4E3J5
GTEX-XBEW-1026
GTEX-XOTO-0526-SM-4B662
GTEX-XPT6-2026-SM-4B64V

GTEX-13FXS-0326
GTEX-13JUV-2326
GTEX-13N11-2726
GTEX-13N2G-2326
GTEX-13NZ9-0626
GTEX-13NZB-2626
GTEX-13O61-2326
GTEX-13OVG-2126
GTEX-13OVH-0626
GTEX-13OVI-1726
GTEX-13OW6-0626
GTEX-13PL7-0626
GTEX-13PVR-2526
GTEX-13QBU-2426
GTEX-13QJ3-0726
GTEX-13S7M-0326
GTEX-13S86-2326
GTEX-13U4I-1826
GTEX-13VXT-0326
GTEX-13W3W-2626
GTEX-13W46-0726
GTEX-13YAN-0526
GTEX-144GL-0326
GTEX-144GM-2026
GTEX-144GN-2426
GTEX-145LT-1626
GTEX-145LV-2326
GTEX-145ME-2026
GTEX-145MI-0326
GTEX-145MN-2426
GTEX-146FH-0526
GTEX-146FQ-0326
GTEX-147F3-0226
GTEX-1497J-2626
GTEX-14A6H-2826
GTEX-14AS3-2126
GTEX-14BMV-0326
GTEX-14C39-2426
GTEX-14ICL-1926
GTEX-O5YT-1626-SM-32PK6
GTEX-OHPK-1626-SM-2YUN3
GTEX-OHPL-1626
GTEX-OHPM-1626
GTEX-XQ8I-0626-SM-4BOPT
GTEX-XUJ4-2626-SM-4BOQ3
GTEX-XUW1-0826-SM-4BOP6
GTEX-XUYS-0326-SM-47JX2
GTEX-XUZC-2126-SM-4BRW8
GTEX-XV7Q-2926-SM-4BRUL
GTEX-XYKS-2426-SM-4AT43
GTEX-Y114-2526
GTEX-Y3IK-2626
GTEX-Y5LM-2126
GTEX-Y5V5-2526
GTEX-Y5V6-2626
GTEX-Y8E4-1026
GTEX-Y8E5-0326
GTEX-Y8LW-2026
GTEX-Y9LG-1926
GTEX-YB5E-2226
GTEX-YB5K-2326
GTEX-YBZK-0326
GTEX-YEC3-2126
GTEX-YEC4-2226
GTEX-YF7O-2526
GTEX-YFC4-1026
GTEX-Z9EW-1726
GTEX-ZA64-2026
GTEX-ZAKK-0326
GTEX-ZC5H-0326
GTEX-ZDYS-1726
GTEX-ZPCL-2026
GTEX-ZPIC-2526
GTEX-ZQG8-1226
GTEX-ZQUD-1726
GTEX-ZT9X-1826
GTEX-ZTPG-0126
GTEX-ZTX8-1626
GTEX-ZV6S-2126
GTEX-ZV7C-2426
GTEX-ZVZO-0326
GTEX-ZVZP-2526
GTEX-ZY6K-2026
GTEX-ZYFG-2426
GTEX-ZYWO-2626
GTEX-ZZ64-1526

**Whole blood**

GTEX-113JC-0006-SM-5O997
GTEX-1192W-0005-SM-5NQBQ
GTEX-11DXX-0005-SM-5NQ8B
GTEX-11EMC-0006-SM-5O9DN
GTEX-11GSP-0006-SM-5N9EL
GTEX-11I78-0005-SM-5N9GB
GTEX-11LCK-0005-SM-5O98U
GTEX-11OF3-0006-SM-5O9CM
GTEX-11ONC-0005-SM-5O9CY
GTEX-11P7K-0006-SM-5N9FM
GTEX-11P82-0006-SM-5N9FY
GTEX-11TT1-0005-SM-5NQ8Y
GTEX-11VI4-0006-SM-5N9D8
GTEX-11WQK-0005-SM-5O9AV
GTEX-11ZTT-0006-SM-5N9FX
GTEX-1212Z-0006-SM-5NQ8M
GTEX-1269C-0005-SM-5N9CJ
GTEX-12C56-0006-SM-5N9E9
GTEX-12KS4-0005-SM-5SI94
GTEX-12WSI-0005-SM-5O99K
GTEX-12WSK-0006-SM-5NQA1
GTEX-12WSM-0005-SM-5NQB3
GTEX-12WSN-0006-SM-5NQAP
GTEX-12ZZX-0005-SM-5O9A9
GTEX-13113-0006-SM-5NQ7X
GTEX-1314G-0005-SM-5NQ9O
GTEX-131XE-0006-SM-5P9F9
GTEX-131XG-0006-SM-5O9CE
GTEX-132NY-0005-SM-5O9AC
GTEX-1399R-0006-SM-5N9FR
GTEX-139UW-0005-SM-5NQ8U
GTEX-13CF3-0006-SM-5N9ED
GTEX-13FTX-0005-SM-5N9F6
GTEX-13FXS-0006-SM-5O99X
GTEX-13OVG-0005-SM-5P9HA
GTEX-13OVH-0005-SM-5P9HB
GTEX-13OVI-0001-SM-5O9BL
GTEX-13OVK-0006-SM-5O9B7
GTEX-13OVL-0006-SM-5O996
GTEX-13OW6-0005-SM-5NQ9Z
GTEX-13OW8-0005-SM-5NQAC
GTEX-13PL7-0005-SM-5N9ET
GTEX-13S7M-0005-SM-5NQ76
GTEX-13VXT-0005-SM-5N9F3
GTEX-147F3-0005-SM-5N9FI
GTEX-147JS-0006-SM-5NQ7K
GTEX-148VI-0006-SM-5O9A6
GTEX-14A5H-0006-SM-5O9AI
GTEX-14AS3-0006-SM-5NQC2
GTEX-14B4R-0006-SM-5O9A7
GTEX-14BMV-0005-SM-5NQ6Y
GTEX-14C38-0006-SM-5NQBF
GTEX-14C39-0005-SM-5NQBR
GTEX-QEG5-0006-SM-2I5FZ
GTEX-QESD-0006-SM-2I5G6
GTEX-R55C-0005-SM-3GAE9
GTEX-RWS6-0005-SM-2XCAN
GTEX-S341-0006-SM-3NM8D
GTEX-SSA3-0005-SM-32QOT
GTEX-T5JW-0005-SM-3GADE
GTEX-T6MN-0005-SM-32PLJ
GTEX-T6MO-0006-SM-32QOU
GTEX-T8EM-0006-SM-3DB71
GTEX-TKQ1-0006-SM-33HBI
GTEX-TKQ2-0006-SM-33HBH
GTEX-TML8-0005-SM-32QPA
GTEX-TMZS-0006-SM-3DB8G
GTEX-U3ZG-0006-SM-47JWX
GTEX-U3ZH-0005-SM-3DB72
GTEX-U4B1-0006-SM-3DB8E
GTEX-UJMC-0005-SM-3GACU
GTEX-UPJH-0006-SM-3GACW
GTEX-V1D1-0006-SM-3NMCE
GTEX-V955-0005-SM-3P5ZC
GTEX-VJYA-0005-SM-3P5ZD
GTEX-VUSG-0006-SM-3GIK9
GTEX-WCDI-0005-SM-3NB2M
GTEX-WFG7-0005-SM-3GIKM
GTEX-WFON-0005-SM-3NMC9
GTEX-WH7G-0005-SM-3NMBX
GTEX-WHPG-0006-SM-3NMBV
GTEX-WHSB-0005-SM-3LK7C
GTEX-WHWD-0005-SM-3LK7D
GTEX-WOFL-0006-SM-3TW8K
GTEX-WOFM-0005-SM-3MJF3
GTEX-WQUQ-0006-SM-3MJF4
GTEX-WRHK-0005-SM-3MJF5
GTEX-WRHU-0006-SM-3MJF6
GTEX-WVLH-0006-SM-3MJF7
GTEX-WXYG-0005-SM-3NB3M
GTEX-WY7C-0006-SM-3NB3L
GTEX-WYVS-0006-SM-3NMA7
GTEX-WZTO-0006-SM-3NM9T
GTEX-X15G-0005-SM-3NMDA
GTEX-X3Y1-0006-SM-3P5ZG
GTEX-X5EB-0006-SM-46MV5
GTEX-X638-0005-SM-47JX6
GTEX-X88G-0006-SM-47JX5
GTEX-XBED-0006-SM-47JXO
GTEX-XBEW-0006-SM-4AT4E
GTEX-XMK1-0005-SM-4B665
GTEX-XPT6-0006-SM-4B66Q
GTEX-XXEK-0005-SM-4BRWJ
GTEX-XYKS-0005-SM-4BRUD
GTEX-Y114-0006-SM-4TT76
GTEX-Y5LM-0005-SM-4V6EJ

GTEX-14DAR-0006-SM-5N9GC
GTEX-14E1K-0006-SM-5N9DY
GTEX-14H4A-0006-SM-5N9E3
GTEX-14ICK-0006-SM-5NQB5
GTEX-14ICL-0006-SM-5SIAB
GTEX-N7MT-0007-SM-3GACQ
GTEX-O5YT-0007-SM-32PK7
GTEX-O5YW-0006-SM-3LK6E
GTEX-OHPL-0006-SM-3MJHB
GTEX-OIZF-0006-SM-2I5GQ
GTEX-OIZI-0005-SM-2XCED
GTEX-OXRP-0006-SM-2I3FN
GTEX-P4QS-0005-SM-2I3EY
GTEX-P78B-0005-SM-2I5GM
GTEX-PLZ5-0006-SM-5S2W5
GTEX-PLZ6-0006-SM-33HBZ
GTEX-POMQ-0006-SM-5SI7D
GTEX-PSDG-0005-SM-3GADC
GTEX-PVOW-0006-SM-3NMB8
GTEX-PW2O-0006-SM-2I3DV
GTEX-PWCY-0005-SM-33HBP
GTEX-Q2AG-0005-SM-5SI7F
GTEX-Q2AH-0005-SM-33HBR
GTEX-Q2AI-0006-SM-2I3FG
GTEX-QCQG-0006-SM-5SI8M
GTEX-Y5V5-0006-SM-4V6FE
GTEX-Y5V6-0005-SM-4V6FD
GTEX-Y8E4-0006-SM-4V6EW
GTEX-Y8E5-0006-SM-47JWQ
GTEX-Y8LW-0005-SM-4V6EV
GTEX-Y9LG-0006-SM-4VBRK
GTEX-YB5K-0005-SM-4VDSP
GTEX-YBZK-0005-SM-59HKG
GTEX-YFC4-0006-SM-4RGLV
GTEX-ZC5H-0005-SM-4WAXM
GTEX-ZDYS-0002-SM-4WKGR
GTEX-ZE9C-0006-SM-4WKG2
GTEX-ZF29-0006-SM-4WKGQ
GTEX-ZGAY-0006-SM-4WWAQ
GTEX-ZP4G-0006-SM-4WWE6
GTEX-ZPIC-0005-SM-4WWEB
GTEX-ZPU1-0006-SM-4WWAT
GTEX-ZQG8-0005-SM-4YCEH
GTEX-ZQUD-0005-SM-4YCE5
GTEX-ZVE2-0006-SM-51MRW
GTEX-ZVP2-0005-SM-51MRK
GTEX-ZVT2-0005-SM-57WBW
GTEX-ZVZP-0006-SM-51MSW
GTEX-ZXES-0005-SM-57WCB

**EBV-transformed lymphocytes (LCLs)**

GTEX-1122O-0003-SM-5Q5DL
GTEX-11EM3-0001-SM-5Q5BD
GTEX-11EMC-0002-SM-5Q5DO
GTEX-11OC5-0004-SM-5S2O6
GTEX-11P7K-0003-SM-5S2OU
GTEX-11TT1-0004-SM-5S2NT
GTEX-11VI4-0001-SM-5S2OI
GTEX-1212Z-0002-SM-5SI6W
GTEX-1269C-0003-SM-5S2PB
GTEX-12BJ1-0003-SM-5SI6V
GTEX-12C56-0002-SM-5S2PC
GTEX-RWS6-0001-SM-3NMAL
GTEX-S4Q7-0003-SM-3NM8M
GTEX-S95S-0002-SM-3NM8K
GTEX-SN8G-0001-SM-3NM8L
GTEX-T5JC-0001-SM-3NMAK
GTEX-T5JW-0003-SM-3NMAD
GTEX-T6MN-0002-SM-3NMAH
GTEX-T6MO-0003-SM-3NMAG
GTEX-TKQ1-0003-SM-3NMAE
GTEX-TML8-0001-SM-3NMAF
GTEX-U3ZH-0002-SM-3NMDD
GTEX-U3ZM-0002-SM-3NMDM
GTEX-U3ZN-0002-SM-3NMDF
GTEX-UPJH-0001-SM-3NMDE
GTEX-UPK5-0003-SM-3NMDI
GTEX-V1D1-0003-SM-3NMDP
GTEX-VJYA-0001-SM-3NMDJ
GTEX-VUSG-0003-SM-3NMDK
GTEX-W5WG-0002-SM-3NMDN
GTEX-W5X1-0001-SM-3P61V
GTEX-WFG7-0001-SM-3P61S
GTEX-WFG8-0001-SM-4LVN8
GTEX-WFJO-0002-SM-3P61X
GTEX-WFON-0001-SM-3P61W
GTEX-WHPG-0004-SM-3NMDO
GTEX-WHSB-0002-SM-4M1ZR
GTEX-WOFM-0001-SM-4OOT2
GTEX-WRHK-0001-SM-4WWDD
GTEX-WWTW-0002-SM-4MVNH
GTEX-WXYG-0004-SM-4MVOS
GTEX-WY7C-0004-SM-4ONDS
GTEX-WYVS-0004-SM-4ONDT
GTEX-WZTO-0001-SM-4PQZY
GTEX-X4LF-0002-SM-4QASG
GTEX-X5EB-0004-SM-46MWA
GTEX-XBED-0003-SM-47JWP
GTEX-XBEW-0002-SM-4AT5O
GTEX-XGQ4-0004-SM-4AT5S
GTEX-XMK1-0001-SM-4B64F
GTEX-XPT6-0001-SM-4B64G
GTEX-XQ3S-0001-SM-4B64K
GTEX-XXEK-0004-SM-4BRWO
GTEX-XYKS-0002-SM-4BRWN
GTEX-Y114-0002-SM-4TT78
GTEX-Y3IK-0001-SM-4WWE1
GTEX-Y5LM-0003-SM-4V6G1
GTEX-Y5V5-0001-SM-4V6FZ
GTEX-Y5V6-0003-SM-4V6FX
GTEX-Y8DK-0004-SM-4RGM7
GTEX-Y8E4-0003-SM-4V6FY
GTEX-Y9LG-0001-SM-4VBRQ
GTEX-YB5E-0001-SM-4VDSV
GTEX-YB5K-0003-SM-4VDSN
GTEX-YEC3-0002-SM-4W1YI
GTEX-YEC4-0002-SM-4W1Z6
GTEX-YF7O-0004-SM-4W1ZT
GTEX-YFCO-0003-SM-4W21I
GTEX-ZC5H-0004-SM-4WAXK
GTEX-ZDTS-0001-SM-4WAXW
GTEX-ZDTT-0004-SM-4WKG3
GTEX-ZEX8-0004-SM-4WKFQ
GTEX-ZF29-0002-SM-4WKF2
GTEX-ZF2S-0004-SM-4WKFE
GTEX-ZF3C-0001-SM-4WWAW
GTEX-ZG7Y-0003-SM-4WWEJ
GTEX-ZLWG-0004-SM-4WWD5
GTEX-ZP4G-0003-SM-4WWED
GTEX-ZPIC-0002-SM-4WWEC
GTEX-ZPU1-0004-SM-4WWAV
GTEX-ZQG8-0001-SM-4YCDH
GTEX-ZQUD-0003-SM-4YCD3
GTEX-ZT9W-0003-SM-4YCE6
GTEX-ZT9X-0004-SM-4YCDT
GTEX-ZTPG-0002-SM-4YCEI
GTEX-ZUA1-0002-SM-4YCF7
GTEX-ZV6S-0003-SM-4YCCT
GTEX-ZV7C-0003-SM-4YCF6
GTEX-ZVT2-0001-SM-57WCK
GTEX-ZVTK-0003-SM-51MRV
GTEX-ZVZP-0004-SM-51MS8

**Cultured fibroblasts**

GTEX-111YS-0008-SM-5Q5BH
GTEX-113JC-0008-SM-5QGR6
GTEX-117XS-0008-SM-5Q5DQ
GTEX-1192W-0008-SM-5QGRE
GTEX-11DXX-0008-SM-5Q5B8
GTEX-11DXY-0008-SM-5QGR4
GTEX-11EMC-0008-SM-5Q5DR
GTEX-11GSP-0008-SM-5Q5DM
GTEX-11I78-0008-SM-5Q5DI
GTEX-11LCK-0008-SM-5Q5BB
GTEX-11NSD-0008-SM-5Q5BC
GTEX-11NUK-0008-SM-5Q5B9
GTEX-11NV4-0008-SM-5Q5BA
GTEX-11O72-0008-SM-5Q5DN
GTEX-11OC5-0008-SM-5S2OH
GTEX-11OF3-0008-SM-5S2NH
GTEX-11ONC-0008-SM-5S2MG
GTEX-11P7K-0008-SM-5S2O5
GTEX-11P81-0008-SM-5S2OT
GTEX-11P82-0008-SM-5S2MS
GTEX-11PRG-0008-SM-5S2N5
GTEX-11TT1-0008-SM-5S2P8
GTEX-11TUW-0008-SM-5SI6S
GTEX-11WQC-0008-SM-5SI6R
GTEX-11WQK-0008-SM-5SI6T
GTEX-11XUK-0008-SM-5S2WD
GTEX-11ZTS-0008-SM-5S2VC
GTEX-11ZTT-0008-SM-5S2TZ
GTEX-11ZUS-0008-SM-5S2UO
GTEX-1211K-0008-SM-5S2W1
GTEX-12126-0008-SM-5S2UC
GTEX-12WSH-0008-SM-5S2V1
GTEX-12WSM-0008-SM-5S2VD
GTEX-1399U-0008-SM-5S2VE
GTEX-N7MS-0008-SM-4E3JI
GTEX-NFK9-0008-SM-4E3JE
GTEX-NL3G-0008-SM-4E3JX
GTEX-O5YT-0008-SM-4E3IQ
GTEX-O5YW-0008-SM-4E3IE
GTEX-OHPK-0008-SM-4E3JL
GTEX-OHPL-0008-SM-4E3I9
GTEX-OHPM-0008-SM-4E3IP
GTEX-OHPN-0008-SM-4E3HW
GTEX-OIZG-0008-SM-4E3J2
GTEX-OIZI-0008-SM-2XCFD
GTEX-OOBJ-0008-SM-3NB26
GTEX-OOBK-0008-SM-3NB27
GTEX-OXRK-0008-SM-3NB28
GTEX-T2IS-0008-SM-4DM75
GTEX-T5JC-0008-SM-4DM6A
GTEX-U4B1-0008-SM-4DXUW
GTEX-U8T8-0008-SM-4DXSP
GTEX-UJHI-0008-SM-4IHL1
GTEX-UJMC-0008-SM-4IHKK
GTEX-UPK5-0008-SM-4IHJD
GTEX-V1D1-0008-SM-4JBIJ
GTEX-W5X1-0008-SM-4LMKA
GTEX-WFG7-0008-SM-4LMKB
GTEX-WHPG-0008-SM-4M1ZQ
GTEX-WHSB-0008-SM-4M1ZP
GTEX-WHWD-0008-SM-4OOSU
GTEX-WI4N-0008-SM-4OOSV
GTEX-WL46-0008-SM-4OOSW
GTEX-WQUQ-0008-SM-4OOT1
GTEX-WRHU-0008-SM-4MVPB
GTEX-WVJS-0008-SM-4MVPC
GTEX-WVLH-0008-SM-4MVPD
GTEX-WY7C-0008-SM-4ONDW
GTEX-WYBS-0008-SM-4ONDX
GTEX-WYJK-0008-SM-4ONDV
GTEX-WYVS-0008-SM-4ONDY
GTEX-WZTO-0008-SM-4PQZZ
GTEX-X15G-0008-SM-4PR2D
GTEX-X3Y1-0008-SM-4PR12
GTEX-X4LF-0008-SM-4QAST
GTEX-XBEC-0008-SM-4AT3X
GTEX-XBEW-0008-SM-4AT3Y
GTEX-XMD2-0008-SM-4WWE7
GTEX-XMD3-0008-SM-4AT4V
GTEX-XMK1-0008-SM-4GICF
GTEX-XOT4-0008-SM-4B664
GTEX-XPT6-0008-SM-4B64Q
GTEX-XPVG-0008-SM-4GICH
GTEX-XQ3S-0008-SM-4GIDZ
GTEX-XUW1-0008-SM-4BOQH
GTEX-XV7Q-0008-SM-4BRWL
GTEX-Y8E4-0008-SM-4V6FW
GTEX-Y9LG-0008-SM-4VBRJ
GTEX-YB5K-0008-SM-4VDT8
GTEX-YEC4-0008-SM-4W1YR
GTEX-YF7O-0008-SM-4W1ZS
GTEX-YJ89-0008-SM-4RGM4
GTEX-Z93S-0008-SM-4RGM5
GTEX-ZC5H-0008-SM-4WAX8
GTEX-ZDTS-0008-SM-4E3I8
GTEX-ZDTT-0008-SM-4E3K5

GTEX-OXRL-0008-SM-3NB29
GTEX-P4PP-0008-SM-48TDV
GTEX-P4QT-0008-SM-48TDZ
GTEX-PSDG-0008-SM-48TE5
GTEX-PW2O-0008-SM-48TEB
GTEX-PWCY-0008-SM-48TE9
GTEX-PX3G-0008-SM-48U2L
GTEX-Q2AH-0008-SM-48U2J
GTEX-QCQG-0008-SM-48U2G
GTEX-QLQ7-0008-SM-447AW
GTEX-QXCU-0008-SM-48FCH
GTEX-R45C-0008-SM-48FF2
GTEX-R55C-0008-SM-48FCF
GTEX-R55D-0008-SM-48FEV
GTEX-R55E-0008-SM-48FCG
GTEX-R55G-0008-SM-48FEX
GTEX-RM2N-0008-SM-48FF3
GTEX-RN64-0008-SM-48FEZ
GTEX-RNOR-0008-SM-48FEY
GTEX-RU1J-0008-SM-46MV9
GTEX-RU72-0008-SM-46MV8
GTEX-RWS6-0008-SM-47JYV
GTEX-RWSA-0008-SM-47JYX
GTEX-S33H-0008-SM-4AD6C
GTEX-S4Z8-0008-SM-33HAZ
GTEX-SE5C-0008-SM-4B64J
GTEX-SJXC-0008-SM-4DM7G
GTEX-ZDXO-0008-SM-4E3HR
GTEX-ZDYS-0008-SM-4E3IX
GTEX-ZE7O-0008-SM-4E3JQ
GTEX-ZEX8-0008-SM-4E3JU
GTEX-ZF2S-0008-SM-4E3IK
GTEX-ZF3C-0008-SM-4E3IL
GTEX-ZLWG-0008-SM-4E3J4
GTEX-ZP4G-0008-SM-4E3I4
GTEX-ZPIC-0008-SM-4E3JF
GTEX-ZPU1-0008-SM-4E3IR
GTEX-ZQG8-0008-SM-4E3J9
GTEX-ZQUD-0008-SM-4YCCU
GTEX-ZT9W-0008-SM-4YCDJ
GTEX-ZT9X-0008-SM-4YCD7
GTEX-ZTPG-0008-SM-4YCEK
GTEX-ZTX8-0008-SM-4YCDV
GTEX-ZUA1-0008-SM-4YCEW
GTEX-ZV68-0008-SM-4YCCV
GTEX-ZV6S-0008-SM-4YCF9
GTEX-ZV7C-0008-SM-57WCL
GTEX-ZVE2-0008-SM-51MRU
GTEX-ZVP2-0008-SM-51MSL
GTEX-ZVT2-0008-SM-57WC9
GTEX-ZVT3-0008-SM-51MRI
GTEX-ZVTK-0008-SM-57WDA
GTEX-ZVZP-0008-SM-51MSX
GTEX-ZXES-0008-SM-57WCX
